# Nominal and real cigarette Price trends from a global dataset, 2008-2024 and implications for the World Health Organization’s 3 by 35 initiative: a longitudinal study and forecasting analysis

**DOI:** 10.64898/2026.09.28.26364093

**Authors:** Rajeev Cherukupalli, Guillermo A. Sandoval, Anne-Marie Perucic, Mark Goodchild, Jeremias Paul

## Abstract

**Introduction:** In July 2025, WHO launched the “3 by 35” Initiative calling on countries to increase real prices of tobacco, alcohol, and sugary drinks by at least 50% by 2035 to secure an additional $ 1 trillion in public revenue. While the effectiveness of excise taxes and prices in reducing tobacco use is well-documented, the feasibility of this call to action has not been evaluated empirically across the world.

**Methods:** Trend changes in countries’ nominal and inflation-adjusted retail prices were investigated using the most-sold brand of cigarettes over windows ranging from 2 to 16 years over the period 2008-2024, derived from nine editions of the World Health Organization Report on the Global Tobacco Epidemic. Statistical models of the association of cigarette excise taxes and prices from the panel of 195 countries were estimated. The frequencies of cigarette prices rising by at least 20% and 50% over preceding periods was estimated, and the length of time countries would need to reach these targets based on past trends were predicted.

**Results:** Over one-fifth of nominal cigarettes prices (and up to 40% in WHO’s African region) were sticky across successive periods, even in countries with high overall consumer price inflation. Between 2008 and 2024, inflation-adjusted international dollar cigarette prices rose at a least squares annual trend growth rate of 2·9% (95% UI 2·8–3·0%) Excise tax increases strongly predict price increases. One-third of countries saw real prices at least 50% higher than 10 years prior.

**Conclusion:** Many of the instances of real price increases of 50% or more over 10-year periods occurred without countries implementing a conscious strategy to raise taxes. It is feasible to achieve and even surpass WHO’s 3 by 35 call to action for tobacco if countries strategize and accelerate tobacco tax increases.

**What is already known:** Previous research has evaluated the impact of cigarette and tobacco taxation and found that prices respond to higher excise taxes. However, whether price increases match, exceed or fall short of general price inflation in a country is an empirical question, with some evidence that taxes are over-shifted in high-income settings and that prices tend to be higher in countries with specific (per unit) taxes. In the policy setting, the Task Force on Fiscal Policy for Health modeled the impact of higher taxes that raise inflation-adjusted prices by at least 50% on reducing the consumption of tobacco. The World Health Organization’s database of cigarette product prices and taxes collected every two years is comprehensive and permits analysis covering many more low- and middle-income countries and years than commercial databases.

**What this study adds:** The study examined the feasibility of WHO’s global call to raise real tobacco prices using all data from available reports of the WHO Report on the Global Tobacco Epidemic from 2008-24. Analysis not previously conducted at this scale (16 years, more than 180 countries) include an examination of the stickiness of nominal cigarette prices, calculation of trends in nominal and real international prices, calculation of the extent of real price increases over multiple time windows, and estimation of country-specific trends real price changes.

**How this study might affect research, practice or policy:** This study finds that nominal cigarette prices were sticky over short time windows in many countries and that globally, real cigarette prices rose on average 2·9% annually. It affirms that excise taxes on cigarettes tend to be over-shifted to prices (that is, a 1 dollar increase in tax increases prices by more than a dollar) especially when specific excise taxes were introduced or increased. One-third of countries saw real cigarette prices rise 50% or more over 10 years, but applying historical trend rates of real price growth based on their 2008-2024 experience indicated that many other countries would fall short of raising prices by 50%. Countries at all income levels have the potential for large cigarette price increases of 50% or more, but will have to accelerate their efforts substantially to achieve the WHO 3 by 35 global call.

## Introduction

In 2025, the World Health Organization (WHO) launched the 3 by 35 Initiative encouraging countries to increase real (inflation-adjusted) prices of tobacco, alcohol, and sugary drinks by at least 50% by 2035. [1–3]. The Initiative builds on the recognition that health taxes are key to reducing the consumption of unhealthy products and reducing deaths from non-communicable diseases. [4–7]. Excise taxes are the primary policy instrument governments use to raise cigarette prices to reduce demand.[8] Multiple studies show they are effective in doing so [9,10]. With the proliferation of data sources on product prices in recent years, economists’ understanding of pricing dynamics in general, and cigarette prices in particular, has grown substantially. [11–14] Prices are termed “sticky” when there are sector- or firm-specific lags in response to underlying cost increases.[15]

Excise taxes, which are a cost to companies, are not always passed on consistently as price increases. Excise taxes may be specific (a fixed amount per cigarette pack), ad valorem (a rate which is multiplied by price), or a combination [8]. Whether, and to what extent, higher cigarette taxes raise prices—not just in nominal terms, but relative to other goods and services—is central to the success of tobacco tax policy to reducing demand[8] The pass-through of excise taxes-i.e., how much a given tax translates to a price increase, is of practical policy interest. A dollar increase in excise taxes may be passed on less thank or more than or exactly as much as a dollar increase in price, i.e. may be undershifted, overshifted, or passed through 1 for 1.The tobacco industry in most countries is highly concentrated and oligopolistic, and companies have a range of price responses when taxes rise. Undershifting implies companies absorb part of a tax increase, usually as a way to keep prices competitive and grow market share. Undershifting may also occur for particular brands and product lines – companies might absorb taxes more to protect the share of a popular or cheap brand. Overshifting, on the other hand, reflects pricing power in the presence of inelastic demand. [8]

Tax targets and calls to action like WHO’s can serve as an important impetus for global tobacco control efforts [16] Historical and country-specific price studies can identify average trends and exemplars to understand the feasibility of achieving targets but are typically limited to a single popular brand, smaller group of countries or the largest global markets covered in commercial databases [17,18]. This study aims to provide new insights on nominal and real cigarette price trend, their relation to excise tax changes, and the occurrence of substantial price increases using a large database of prices and taxes (over 180 countries) systematically collected every two years since 2008.

## Methods

Our study combined a retrospective analysis of nominal and real price trends, a statistical evaluation of the pass-through from taxes to prices, and forecasts of when countries would reach higher real prices corresponding to WHO’s call.

### Data sources

Data on nominal and real prices was assembled from nine editions of the WHO Report on the Global Tobacco Epidemic over the period 2008-24. [19,20] WHO collects detailed information on taxes applied to tobacco products (cigarettes and other combustible and non-combustible products) from 195 Member States every two years. The data is validated, draws on a standardized survey instrument and has been used since 2008 as the basis for technical outreach and additional cross-country analysis and data presentations in the field of tobacco control.

Information on the most-sold brand is obtained independently when possible, and questionnaires are pre-filled with the brand, with respondents having the option of reporting a different brand if needed. Data on prices of the most-sold brand are collected by WHO through regional and country focal points, by surveying stores (kiosks, supermarkets), typically in the capital of the country. Prices are also compared, when possible, to online sources. In each round of the exercise, information on the previous round’s taxes and tax calculations (and prices, in rarer instances) is updated if discrepancies are found or if countries report corrections.

WHO records prices in the local currency used as on July 31 of the year of data collection. In the case of currency changes, WHO also reports a consistent series of nominal prices by converting the older currency to the newer one adopted. WHO calculates and reports real local currency prices using annual consumer price inflation series from the World Bank.[21] Prices are also expressed in international dollars using purchasing power parity conversion factors sourced from the IMF’s World Economic Outlook database.[22]

The reliance on the most-sold brand to represent the market is partly a matter of convenience but also reflects the relatively high concentration of the tobacco industry, where a single brand can account for as much as 50% or more of the market share. [20] The most-sold brand (i.e. statistical mode) can be different from the average price. While some countries, particularly those in the European Union, are legally obligated to collect and report a weighted average price computed as the value of cigarette sales divided by the volume, governments of most countries, however, do not report aggregate sales values and volume. This is less of a concern in practice, since the data collection aims to obtain a single, easily sourced price measure. Exceptions to using the most-sold brand are Canada, the United States (both with varying sub-national taxes) and China (to summarize a very wide range of brands and price points).WHO-collected prices correlate highly with average weighted prices estimated from globally reported sales data, indicating that the price of the most-sold brand serves as a good approximation of the actual average price in a country.[20]

The dataset includes details on brands. We matched brands at the country level over successive periods, with 87% of price pairs determined to be identical in adjacent periods (supplemental table S1). However, only 63 countries reported an identical brand in each of the 9 periods from 2008 to 2024. The dataset also includes details of the excise taxes, including the type of tax systems, statutory rates and the computed share of excise in retail price (“excise yield”). In computing trend growth rates, countries were dropped if they did not have international prices in at least 4 years, or when inflation was extreme (e.g. Venezuela), reducing the sample to 181 countries

### Statistical analysis

We first examined the nominal and real local and international dollar price behavior of the most-sold brand of cigarettes from using descriptive statistics and tabulations. We estimated the fraction of countries with substantial (20% and 50%) real price increases over successive time windows. We analyzed regional and income-group patterns in pricing, and whether real price increases were likelier when prices were lower to begin with.

Since excise taxes are the primary policy tool to influence cigarette prices, we examined whether higher excise tax increases are reliably associated with higher price increases, first through tabulations of percentage price change by quantiles of tax increase, and then by estimating the pass-through of excise tax to prices in the panel using a two-way fixed effects model. Following Dauchy and Filby (2025), we implemented a simple model of the pass-through of the excise tax (measured in international dollars) to price i.e.

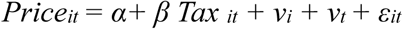

where *Price_it_* and *Tax _it_* represent price and tax on 20 sticks of the most-sold brand measured in international $ for country i in year t, *ν_i_* and *v_t_* and represent country and year fixed effects respectively. [23] The two-way fixed effects model controlled for time- and country-specific trends; standard errors were clustered at the country level to account for the potential correlation within-country.

We estimated the model in levels and in natural logs, with additional time-varying controls for the type of tax system (ad valorem, specific, mixed).

Finally, we estimate the least squares growth rate of real price for each country to calculate the time that each country would need to see real cigarette prices rise 20% and 50% higher and evaluate countries’ preparedness to achieve the WHO 3 by 35 call for tobacco.

### Patient and public involvement statement

No patients or members of the public were involved in this research.

## Results

### Country-level nominal price trends in local currency

22% of prices in the sample were identical to price in the previous period (Table 1). Prices were particularly sticky in the AFRO and EMRO regions (40% and 27%, respectively). Price increases were more frequent in the EURO region, where over 60% of countries recorded prices changing every period (supplemental table S2). At the other extreme, prices remained unchanged in five or more periods in some countries, mostly in west Africa. Sticky nominal prices persist when analysis was restricted to identical brands in successive periods and to identical brands in each of the nine periods (supplemental tables S3 and S4)

**Table 1:** Nominal price stickiness: proportion of most-sold brand local currency prices unchanged from the preceding two-year period.

| Region | 2010 | 2012 | 2014 | 2016 | 2018 | 2020 | 2022 | 2024 | Total |
| --- | --- | --- | --- | --- | --- | --- | --- | --- | --- |
| AFRO | 0.48 | 0.55 | 0.37 | 0.39 | 0.46 | 0.34 | 0.45 | 0.27 | **0.41** |
| AMRO | 0.24 | 0.09 | 0.15 | 0.24 | 0.09 | 0.10 | 0.07 | 0.00 | **0.12** |
| EMRO | 0.36 | 0.14 | 0.20 | 0.35 | 0.33 | 0.24 | 0.28 | 0.29 | **0.27** |
| EURO | 0.10 | 0.02 | 0.04 | 0.12 | 0.02 | 0.10 | 0.15 | 0.04 | **0.07** |
| SEARO | 0.11 | 0.33 | 0.22 | 0.00 | 0.00 | 0.40 | 0.27 | 0.20 | **0.19** |
| WPRO | 0.46 | 0.22 | 0.12 | 0.08 | 0.35 | 0.33 | 0.39 | 0.22 | **0.27** |
| **Total** | **0.30** | **0.21** | **0.18** | **0.21** | **0.21** | **0.22** | **0.26** | **0.14** | **0.22** |

Note: The value in each cell is the fraction of countries in the WHO region reporting nominal prices for the most-sold brand identical to the price in the previous edition of the Report on the Global Tobacco Epidemic; 2010 is in comparison to 2008. Countries had to report prices in both periods to be included. As a result, between 176 and 182 of the 195 countries were included in any given year.

### Aggregate price trends in international dollars

To examine global and regional price trends, local currency prices were converted to international dollars using country- and year-specific purchasing power parity conversion factors, after which they were converted to “real”, i.e. 2024-denominated international dollars using annual CPI inflation for the US. Average prices were highest in the EURO region and high-income countries (supplemental tables S5-S8). The least squares annual trend growth rate of real international dollar cigarette prices was 2·9%(2·8-3·0%) (Table 2). Regional growth rates ranged from 1·5%(1·3-1·7%) to 3·8%(3·6-4·0%) for the AFRO and EURO regions, respectively.

**Table 2:**
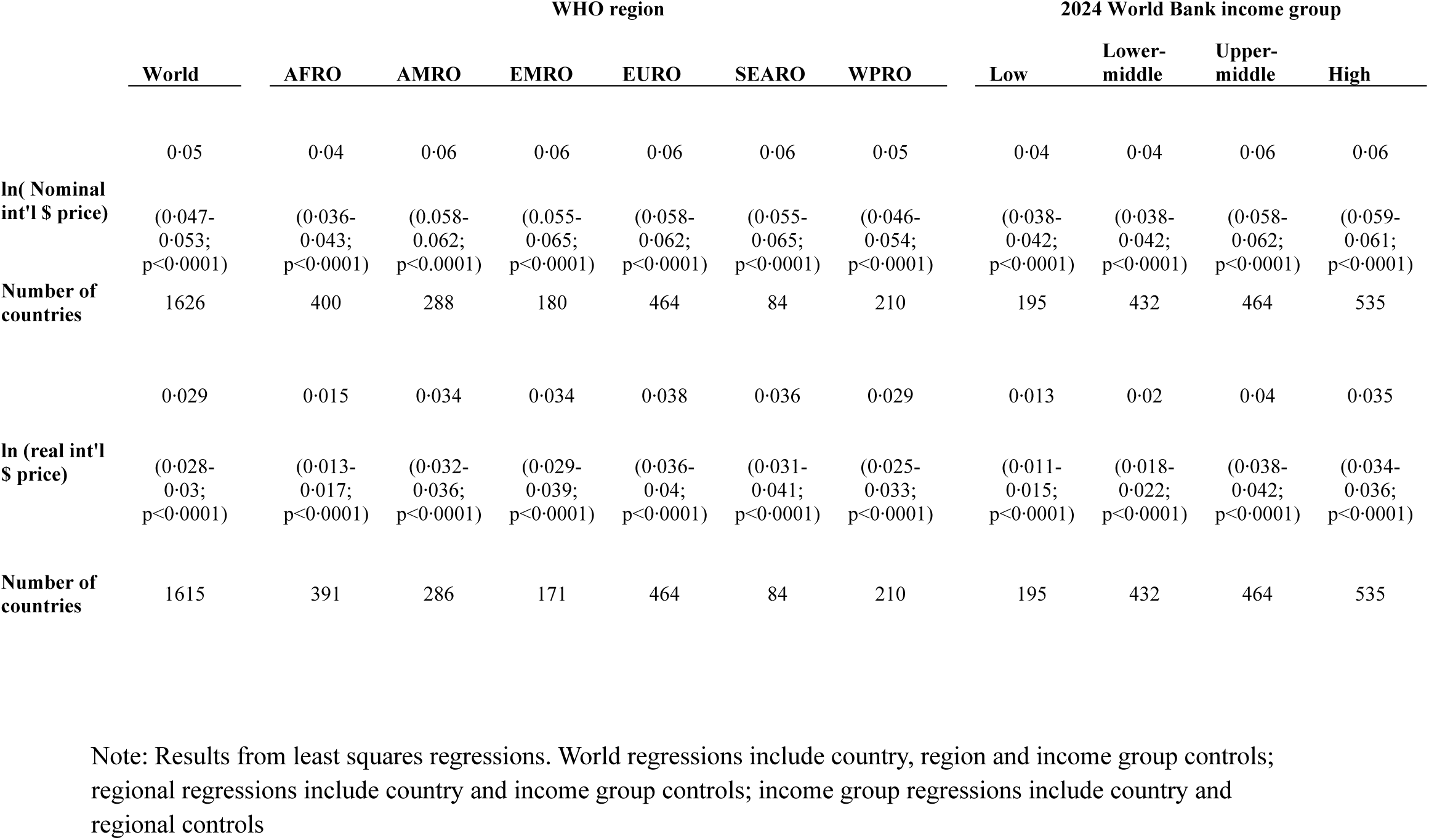
Least squares aggregate trends in international most-sold brand cigarette prices.

|  | WHO region |  |  |  |  |  |  | 2024 World Bank income group |  |  |  |
| --- | --- | --- | --- | --- | --- | --- | --- | --- | --- | --- | --- |
|  | World | AFRO | AMRO | EMRO | EURO | SEARO | WPRO | Low | Lower-middle | Upper-middle | High |
| **ln( Nominal int'l \$ price)** | 0.05<br>(0.047-0.053;<br>p<0.0001) | 0.04<br>(0.036-0.043;<br>p<0.0001) | 0.06<br>(0.058-0.062;<br>p<0.0001) | 0.06<br>(0.055-0.065;<br>p<0.0001) | 0.06<br>(0.058-0.062;<br>p<0.0001) | 0.06<br>(0.055-0.065;<br>p<0.0001) | 0.05<br>(0.046-0.054;<br>p<0.0001) | 0.04<br>(0.038-0.042;<br>p<0.0001) | 0.04<br>(0.038-0.042;<br>p<0.0001) | 0.06<br>(0.058-0.062;<br>p<0.0001) | 0.06<br>(0.059-0.061;<br>p<0.0001) |
| **Number of countries** | 1626 | 400 | 288 | 180 | 464 | 84 | 210 | 195 | 432 | 464 | 535 |
| **ln (real int'l \$ price)** | 0.029<br>(0.028-0.03;<br>p<0.0001) | 0.015<br>(0.013-0.017;<br>p<0.0001) | 0.034<br>(0.032-0.036;<br>p<0.0001) | 0.034<br>(0.029-0.039;<br>p<0.0001) | 0.038<br>(0.036-0.04;<br>p<0.0001) | 0.036<br>(0.031-0.041;<br>p<0.0001) | 0.029<br>(0.025-0.033;<br>p<0.0001) | 0.013<br>(0.011-0.015;<br>p<0.0001) | 0.02<br>(0.018-0.022;<br>p<0.0001) | 0.04<br>(0.038-0.042;<br>p<0.0001) | 0.035<br>(0.034-0.036;<br>p<0.0001) |
| **Number of countries** | 1615 | 391 | 286 | 171 | 464 | 84 | 210 | 195 | 432 | 464 | 535 |

Note: Results from least squares regressions. World regressions include country, region and income group controls; regional regressions include country and income group controls; income group regressions include country and regional controls

### Country-level patterns in inflation-adjusted local currency prices

International dollar prices are generally better suited for cross-country comparisons at a point in time. Over time, international dollar price increases reflect both local real prices and US (rather than country-specific) inflation over time. Median local annual consumer price inflation across the period 2008-2024 was 3.6%, with low- and lower middle-income countries and the AFRO and SEARO region experiencing higher rates (supplemental table S9). Real prices in the local currency at the country-level rarely trend in a smooth manner over time and are often subject to declines and even negative changes (supplemental figure F1). Negative real price changes were fairly common when prices in a given period are compared to the immediately preceding period (supplemental figure F2). While median and mean real prices were only 7% and 3% higher over a 2-year window, they were 77% and 48% higher than 16 years prior (supplemental table S10). The relationship held broadly when comparisons were limited to countries which had real price data in every period and to countries with identical brands in all periods (supplemental tables S11 and S12).

### Incidence of substantial real price increases (20% and higher, 50% and higher)

The Task Force on Fiscal Policy and Health (2019, 2024) models scenarios of tax revenue increase and consumption reductions when all countries implement higher taxes that immediately raise real prices by 20% and 50%. [24–26] WHO’s 3 by 35 Initiative calls for tax increases that raise real prices of any or all of three products (tobacco, alcohol and sugary drinks) by at least 50% by 2035 (a 10-year period).

Figure 1 depicts the fraction of countries where prices were at least 20% and 50% higher relative to prices in each of 8 possible prior periods. After 10 years, real prices rose by at least 20% in 66% of countries, and by at least 50% in 33% of countries. The EURO and AMRO regions, and upper-middle and high-income countries registered the highest incidence of appreciable real price increases (supplemental tables S13 and S14). Countries at the upper end of the world cigarette price distribution were better raising prices over 10 years, with 89% and 55% of countries in the topmost quintile attaining real price increases of over 20% and 50%, respectively (supplemental figure F3).

### Trend growth rates of real prices and time to attain 3 by 35 real price increases

We computed country-specific least squares trend growth rates of real prices, tested for the statistical significance of those trends, and applied the estimated annual growth rate of real prices to project by which year real prices would rise to over 20% and over 50% of their 2024 level (supplemental table S15). At historical rates of price increase, the majority of countries require more than 5 years for real prices to rise over 20%, and more than 10 years for real prices to rise over 50%. Only 51 of 181 countries would achieve a 50% real price increase by 2035 if their past trends continued.

### Pass through from excise taxes to price

Larger excise tax increases (in deciles) and purely specific tax regimes were associated with larger price increases (supplemental tables S16 and S17). We examined instances of real prices rising over 50% over two years and whether they corresponded to a noteworthy excise tax policy change (supplemental table S189). Introducing or increasing a specific tax was the most common reason for substantial two-year increases in real price (29 of 48 instances, supplemental table S19).

Table 3 presents the results of two-stage fixed effects regressions of prices on the excise taxes, both measured in international dollars. We restricted our sample to instances of price change larger than -200% (to account for a few outliers). Across specifications with and without controls for the type of tax system (Columns I and II), a one international dollar increase in excise tax was associated on average with a 1·46(95% CI 1·36−1·56) international dollar increase in price, comparable to Dauchy and Filby’s estimate for cigarettes (1·33 to 1·38 across models). However, unlike Dauchy and Filby, we did not find evidence of undershifting in low- and middle-income countries as a group (Columns III and IV). We repeated the regressions in natural logs and found that 1% increase in excise taxes is associated with a 0·33% (95% CI 0.29−0.37%) increase in price in a specification that included controls for tax regime type (supplemental table S20, column II). Interactions of the excise tax with type of tax regime were not statistically significant (supplemental table S21)

**Table 3:** Pass through in levels: Coefficients from two-way fixed-effects regression of price of cigarettes on excise tax (international $)

|  | Full sample |  | High income, 2024 | LMIC, 2024 |
| --- | --- | --- | --- | --- |
|  | (I) | (II) | (III) | (IV) |
| Excise tax, PPP\$ | 1.46<br>(1.36-1.56;<br>p<0.0001) | 1.46<br>(1.36-1.56;<br>p<0.0001) | 1.25<br>(1.21-1.29;<br>p<0.0001) | 1.66<br>(1.47-1.85<br>p<0.0001) |
| Constant | 1.42<br>(1.20-1.64;<br>pp<0.0001) | 1.22<br>(0.79-1.65;<br>p=0.005) | 1.53<br>(1.40-1.66;<br><0.0001) | 1.47<br>(1.19-1.75;<br>p<0.0001) |
| Mixed |  | 0.05<br>(-0.97-0.33;<br>p=0.845) |  |  |
| Specific |  | 0.51<br>(-0.1-1.04;<br>p=0.331) |  |  |
| Overall R-squared | 0.76 | 0.75 | 0.88 | 0.68 |
| # countries | 181 | 181 | 59 | 122 |
| N (observations) | 1626 | 1626 | 481 | 1004 |

Note: Coefficients from a two-way fixed effects regression of cigarette prices on excise tax yield in international (PPP-adjusted) dollars. Panel with observations in even-numbered years, 2008-24. Robust standard errors (clustered at the country level).

Omitted tax regime category in column II = ad valorem. Restricted to observations where 2-year price changes were greater than -500%n

## Discussion

Taxes on tobacco tend to be among the highest indirect taxes as a fraction of product price, reflecting the inelastic demand for tobacco. Prices in the real world, however, vary widely, as do the rates at which these prices change over time. Intuitively, countries that start in a similar position, including income group and general product price level, can end up with very different price trajectories in the face of overall inflation and industry pricing practices.

Our study has several strengths and practical implications. It is, to our knowledge, the first to document how widespread nominal cigarette price stickiness over the two-to-four-year window is in the world, particularly in low-income countries, a finding which is in line with a large and growing economics literature on price rigidity. We use comparable prices (international dollars) across the entirety of WHO’s database to estimate the trend growth of global, regional and income-group cigarette prices, and find that real prices have risen at less than 3% on average. We offer simple country-specific projections of how quickly countries can achieve large real price increases at current trends as a way for countries to assess their prospects to achieving the 3 by 35 goals in the status quo. We also establish that some of the largest price increases have occurred when countries introduce or increase specific excise taxes.

Sticky nominal prices in the face of inflation suggests that companies prioritize maintaining a particular price point for their largest selling brand, possibly to deter competition. Unchanged prices over the long term need not be unprofitable, when considering the tendency of prices to be in multiples of 10 and 100 units in countries with a history of higher inflation. Some 37% of price observations in the data, ranging from 13% in the EURO region to 77% in AFRO countries, were in exact multiples of 10 (supplemental table S22). When prices are commonly found in multiples of 100 (e.g. 500 XAF, 400 Kenyan shillings, 3500 Guaraní paraguayo), a small percentage increases in costs or taxes can be absorbed without raising prices for a few years if eventually prices are raised in increments of 100 units. Unchanged prices over multiple years are an easily identified phenomenon, and one that merits regulators examining the market structure and tax environment that keep prices low and constant.

Our findings on tax pass-through benefit from using a larger dataset and more years than typically available to confirm that taxes tend to be overshifted to cigarette prices. Unlike other studies, we do not, however, find evidence of undershifting in low- and middle-income countries in this sample. If broadly true, this would suggest that a dollar increase in excise tax increases can reliably raise prices by more than their prices by more than a dollar across settings. We also document that large real price increases of over 50% over the short term (two-years) are possible, particularly when a specific tax is introduced or increased substantially, and when countries with no cigarettes excise introduce one. The few instances when similarly large price increases occurred without a tax increase are a missed opportunity for taxation.

While models of tax-induced price increases assume real cigarette prices rise in a predictable manner (e.g. a large tax induces a large one-time price increase, with real prices staying high thereafter), our study points to the reality of countries’ experiences of price increases being far from smooth on a year-to-year basis, even as most countries do see real prices rise over 10 years and longer. This points to the need for countries to have both short and longer period price surveillance.

Absent a conscious effort to raise taxes, many countries, particularly in the AFRO region risk not seeing substantial real price increases by 2035. By the same measure, small changes can accelerate the process-moving from an annual real price increase of 4.2% to 6.1% reduces the time to raise prices by 50% from 10 to 7 years. And even larger tax and real price increases are possible over shorter periods. In the status quo, 50 countries are on track to raise real prices by 50% or more within 10 years. That count rises to 102 countries if all countries raise real prices at an additional 2% annually, and 181 if countries are able to use tax increases to raise real prices at 6–8% annually, corresponding to the 90^th^ percentile of real price growth rates within their WHO region, (Figure 2).

Our study also has limitations. Relying on one price measure (most-sold brand price) does not capture the entirety of price responses, in particular whether prices in other segments rise faster or slower over time. While we find that higher excise taxes are reliably passed on to higher prices, we are agnostic about the specifics of what drives this relation. Excise tax policy—in particular, the type, complexity, level, frequency of increase and indexation to overall inflation— has a key role in explaining price increases. This link and the sequencing from tax to price increases is directly apparent in the data in the case of individual countries like the Philippines, as is the link from periods when both taxes and nominal prices are unchanged. A more detailed analysis of country tax policy developments and their impact on pricing is beyond the scope of the current exercise.

## Conclusions

WHO’s 3 by 35 initiative calls on countries to use taxes to raise real prices on at least one product (tobacco, alcohol, sugary drinks) by at least 50% by 2035. Using data collected by WHO over 2008-2024, we found that one-third, and up to 40% of countries have seen real cigarette prices rise by 50% or more over the prior 10 years. This progress occurred even as at most 41 countries in any given year achieved WHO’s recommended tobacco taxation of 75% or more of retail price. [20] The 2025 Sevilla Commitment includes a call to increase taxes on tobacco “as a non-distortionary source with a clear potential to increase domestic revenue and reduce the risk factors of noncommunicable diseases.” [27] A conscious push to raise health taxes is therefore likely to lead many more countries to successfully raise the real price of health-harming products both substantively and sooner than 2035.

## Supporting information

Cherukupalli 3 by 35 supplement

## Data Availability

Data sharing
Cigarette price and tax data used in this study is publicly available at WHOs Global Health Observatory (https://www.who.int/data/gho), with the exception of cigarette brand names. Consumer price inflation data is publicly available from the World Bank(https://datatopics.worldbank.org/world-development-indicators). Purchasing power parity conversion factors are available from the International Monetary Fund (https://data.imf.org/en/datasets/IMF.RES:WEO).

https://www.who.int/data/gho

## Contributors

RC designed the study, conducted statistical analysis, interpreted the results, drafted the manuscript and is the guarantor of this work. RC and AMP accessed and verified the data and prepared and cleaned the dataset. GS performed initial tabulations, analysis and drafted an analytical note to support the real price increase targets proposed by the WHO 3 by 35 initiative. RC, GS, MG and JP reviewed and edited the manuscript. All authors had full access to all the data in the study and had final responsibility for the decision to submit for publication.

## Acknowledgments

This research was funded by the World Health Organization through a grant from Bloomberg Philanthropies (Bloomberg.org).

## Data sharing

Cigarette price and tax data used in this study is publicly available at WHO’s Global Health Observatory (https://www.who.int/data/gho), with the exception of cigarette brand names. Consumer price inflation data is publicly available from the World Bank(https://datatopics.worldbank.org/world-development-indicators). Purchasing power parity conversion factors are available from the International Monetary Fund (https://data.imf.org/en/datasets/IMF.RES:WEO).

