## Supplementary material for "Nominal and real cigarette Price trends from a global dataset, 2008-2024 and implications for the World Health Organization’s 3 by 35 initiative: a longitudinal study and forecasting analysis": Cherukupalli 3 by 35 supplement

### Nominal and Real Cigarette Price Trends and Excise Tax Pass-Through: Longitudinal Evidence from a global dataset, 2008-2024 and implications for the World Health Organization's 3 by 35 Initiative

#### Appendix Tables and Figures

Supplemental Table S18: Excise tax changes associated with 2-year PPP$ price increases of over 50%: Instances of two-year increase over 50% and explanations ... 20

Supplemental Table S20: Pass-through of excise tax to prices: Coefficients from two-way fixed-effects regression of ln(price of cigarettes in international $) on ln(Excise Tax in international $) ... 23

Supplemental Table S21: Interaction of price measures and tax regime ... 24

Supplemental Table S22:Terminal digits in nominal local currency prices ... 25

Supplemental Figure F1: Change in real local currency prices over previous period by WHO region (countries with 4 or more price observations) ... 26

Supplemental Figure F2: Distribution of real price changes over 2- and 10-year lookback period ... 29

Supplemental Figure F3: Fraction of countries with real prices 20% and 50% higher by price quintile ... 29

**Supplemental Table S1: Brand matches over successive periods**

| | 2010 | 2012 | 2014 | 2016 | 2018 | 2020 | 2022 | 2024 | Total | Share |
|---|---|---|---|---|---|---|---|---|---|---|
| Initial match, identical brands | 125 | 123 | 162 | 157 | 139 | 148 | 153 | 167 | 1174 | 81% |
| Additional matches | 21 | 20 | 13 | 9 | 8 | 4 | 7 | 4 | 86 | 6% |
| Unmatched | 34 | 37 | 7 | 16 | 35 | 24 | 17 | 12 | 182 | 13% |
| Total # of prices | **180** | **180** | **182** | **182** | **182** | **176** | **177** | **183** | **1442** | 100% |

Brands were matched as follows. First, brands were matched if they were spelled identically (1174 pairs of prices). Additional matches were then obtained. Any small spelling differences, capitalizations and blank spaces in brand names were tallied and corrected (e.g. "Kentuchy" and "Kentucky"). After that brand variants were evaluated and coded as the same brand if they were essentially identical (e.g. Gold Leaf and JP Gold Leaf) or sufficiently similar (e.g., 20 Mars and 20 Mars inter légère), or if multiple brands were provided by a country, with at least one brand overlapping. Marlboro Light and Marlboro (in Israel) and Marlboro and Philip Morris (in Andorra) were coded as the same brand. This process yielded 86 additional matches.

**Supplemental Table S2: Frequency of nominal price stickiness**

Number of countries by count of periods with sticky prices by WHO region (0= prices change in every period. 8 = prices identical in successive periods over 2008-24)

| Region | **0** | **1** | **2** | **3** | **4** | **5** | **6** | **7** | **8** | **Total** |
|---|---|---|---|---|---|---|---|---|---|---|
| AFRO | 4<br>(10.81%) | 3<br>(8.11%) | 8<br>(21.62%) | 4<br>(10.81%) | 7<br>(18.92%) | 4<br>(10.81%) | 1<br>(2.7%) | 3<br>(8.11%) | 3<br>(8.11%) | **37**<br>(100%) |
| AMRO | 16<br>(55.17%) | 6<br>(20.69%) | 3<br>(10.34%) | 2<br>(6.9%) | 2<br>(6.9%) | | | | | **29**<br>(100%) |
| EMRO | 3<br>(17.65%) | 6<br>(35.29%) | 3<br>(17.65%) | 1<br>(5.88%) | 2<br>(11.76%) | 1<br>(5.88%) | 1<br>(5.88%) | | | **17**<br>(100%) |
| EURO | 30<br>(61.22%) | 14<br>(28.57%) | 3<br>(6.12%) | 1<br>(2.04%) | 1<br>(2.04%) | | | | | **49**<br>(100%) |
| SEARO | 3<br>(33.33%) | 1<br>(11.11%) | 3<br>(33.33%) | 2<br>(22.22%) | | | | | | **9**<br>(100%) |
| WPRO | 4<br>(22.22%) | 3<br>(16.67%) | 3<br>(16.67%) | 5<br>(27.78%) | 2<br>(11.11%) | | | 1<br>(5.56%) | | **18**<br>(100%) |
| **Total** | **60**<br>(37.74%) | **33**<br>(20.75%) | **23**<br>(14.47%) | **15**<br>(9.43%) | **14**<br>(8.81%) | **5**<br>(3.14%) | **2**<br>(1.26%) | **4**<br>(2.52%) | **3**<br>(1.89%) | **159**<br>(100%) |

Note: Analysis restricted to countries with a price observation in every time period. Zimbabwe dropped due to multiple currency changes. Figures in parentheses add up to 100 for each region.

**Supplemental Table S3: Nominal price stickiness: share of identical local prices reported over prior period, sample restricted to instances with brands identical to previous year's brand**

| Region | **2010** | **2012** | **2014** | **2016** | **2018** | **2020** | **2022** | **2024** | **Total** |
|---|---|---|---|---|---|---|---|---|---|
| AFRO | 0.50 | 0.58 | 0.39 | 0.42 | 0.44 | 0.32 | 0.44 | 0.30 | **0.43** |
| AMRO | 0.30 | 0.10 | 0.16 | 0.24 | 0.09 | 0.14 | 0.08 | 0.00 | **0.14** |
| EMRO | 0.33 | 0.14 | 0.20 | 0.37 | 0.39 | 0.22 | 0.27 | 0.33 | **0.28** |
| EURO | 0.10 | 0.05 | 0.04 | 0.13 | 0.00 | 0.08 | 0.14 | 0.04 | **0.07** |
| SEARO | 0.14 | 0.38 | 0.22 | 0.00 | 0.00 | 0.40 | 0.27 | 0.20 | **0.21** |
| WPRO | 0.45 | 0.24 | 0.14 | 0.10 | 0.36 | 0.44 | 0.43 | 0.23 | **0.30** |
| **Total** | **0.33** | **0.27** | **0.18** | **0.23** | **0.22** | **0.23** | **0.26** | **0.15** | **0.23** |

Note: The value in each cell is the fraction of countries in the WHO region reporting nominal prices for the most-sold brand identical to the price in the previous edition of the Report on the Global Tobacco Epidemic; 2010 is in comparison to 2008. Countries had to report prices in both periods AND report the same brand as in the previous period to be included. As a result, between 142 and 169 of the 195 countries were included in any given year (vs 176 to 182 countries when previous periods' brands are not matched)

**Supplemental Table S4: Nominal price stickiness: share of identical local prices reported over prior period, sample restricted instances with brands identical in all 9 periods**

| Region | **2010** | **2012** | **2014** | **2016** | **2018** | **2020** | **2022** | **2024** | **Total** |
|---|---|---|---|---|---|---|---|---|---|
| AFRO | 0.39 | 0.53 | 0.47 | 0.47 | 0.35 | 0.24 | 0.47 | 0.22 | **0.39** |
| AMRO | 0.00 | 0.11 | 0.22 | 0.00 | 0.11 | 0.11 | 0.00 | 0.00 | **0.07** |
| EMRO | 0.22 | 0.00 | 0.22 | 0.44 | 0.22 | 0.22 | 0.44 | 0.56 | **0.29** |
| EURO | 0.10 | 0.00 | 0.10 | 0.10 | 0.00 | 0.00 | 0.00 | 0.00 | **0.04** |
| SEARO | 0.20 | 0.00 | 0.00 | 0.00 | 0.00 | 0.20 | 0.40 | 0.40 | **0.15** |
| WPRO | 0.44 | 0.33 | 0.11 | 0.22 | 0.33 | 0.44 | 0.44 | 0.11 | **0.31** |
| **Total** | **0.25** | **0.22** | **0.24** | **0.25** | **0.20** | **0.20** | **0.31** | **0.20** | **0.23** |

Note: The value in each cell is the fraction of countries in the WHO region reporting nominal prices for the most-sold brand identical to the price in the previous edition of the Report on the Global Tobacco Epidemic; 2010 is in comparison to 2008. Countries had to report prices in both periods AND reporting the same brand as all periods 2008-24. As a result, between 59 and 60 of the 195 countries were included in any given year (vs between 142 and 169 countries when only the previous period's brands are matched)

**Supplemental Table S5: Mean real international dollar prices over time across WHO regions**

| | AFRO | AMRO | EMRO | EURO | SEARO | WPRO | All regions |
|---|---|---|---|---|---|---|---|
| **2008** | 4.27 | 5.14 | 3.51 | 5.97 | 6.13 | 5.92 | 5.11 |
| | (2.81) | (2.44) | (1.83) | (2.75) | (3.18) | (3.93) | (2.94) |
| **2010** | 4.37 | 5.80 | 4.04 | 6.56 | 5.98 | 5.91 | 5.49 |
| | (2.73) | (3.04) | (2.40) | (3.02) | (3.56) | (4.00) | (3.16) |
| **2012** | 4.10 | 6.09 | 4.25 | 7.28 | 6.44 | 6.70 | 5.84 |
| | (2.63) | (2.99) | (2.35) | (3.11) | (4.05) | (4.38) | (3.38) |
| **2014** | 4.61 | 6.48 | 4.83 | 8.06 | 6.87 | 6.85 | 6.36 |
| | (2.83) | (3.14) | (2.73) | (3.20) | (4.75) | (4.94) | (3.64) |
| **2016** | 5.07 | 7.25 | 5.39 | 8.84 | 8.83 | 8.25 | 7.19 |
| | (4.07) | (3.60) | (3.04) | (3.40) | (7.48) | (5.79) | (4.43) |
| **2018** | 5.18 | 7.60 | 6.19 | 9.19 | 9.53 | 8.37 | 7.51 |
| | (4.41) | (3.90) | (4.60) | (3.70) | (7.33) | (5.84) | (4.78) |
| **2020** | 5.26 | 8.83 | 7.28 | 10.15 | 11.94 | 9.69 | 8.46 |
| | (3.40) | (4.13) | (5.29) | (4.01) | (8.06) | (7.31) | (5.25) |
| **2022** | 4.98 | 8.12 | 6.80 | 9.92 | 10.66 | 9.14 | 8.04 |
| | (3.15) | (3.67) | (4.16) | (5.03) | (6.85) | (6.78) | (5.08) |
| **2024** | 5.13 | 8.50 | 6.33 | 10.30 | 10.67 | 9.69 | 8.23 |
| | (3.22) | (3.59) | (4.45) | (5.14) | (9.04) | (7.43) | (5.43) |

Calculated for countries with price data in real international dollar price data (177 to 181 countries over 9 periods)

**Supplemental Table S6: Median real international dollar prices over time across WHO regions**

| | AFRO | AMRO | EMRO | EURO | SEARO | WPRO | All regions |
|---|---|---|---|---|---|---|---|
| **2008** | 3.41 | 4.68 | 3.13 | 5.92 | 5.83 | 5.55 | 4.53 |
| **2010** | 3.73 | 5.15 | 3.74 | 6.71 | 6.02 | 4.88 | 4.89 |
| **2012** | 3.08 | 5.86 | 3.85 | 7.69 | 5.56 | 5.61 | 5.25 |
| **2014** | 3.36 | 5.70 | 4.64 | 8.34 | 6.52 | 5.36 | 5.42 |
| **2016** | 3.65 | 6.40 | 5.45 | 8.90 | 7.54 | 7.10 | 6.41 |
| **2018** | 3.65 | 7.02 | 5.31 | 9.16 | 7.62 | 6.82 | 6.66 |
| **2020** | 4.14 | 8.15 | 5.56 | 9.60 | 9.91 | 6.86 | 7.60 |
| **2022** | 3.96 | 7.64 | 6.31 | 9.13 | 8.72 | 7.27 | 7.18 |
| **2024** | 4.07 | 7.78 | 4.59 | 9.21 | 8.86 | 7.37 | 7.44 |

Calculated for countries with price data in real international dollar price data (177 to 181 countries over 9 periods)

**Supplemental Table S7: Mean real international dollar prices over time across World Bank 2024 income groups**

| | Low | Lower-middle | Upper-middle | High | All countries |
|---|---|---|---|---|---|
| **2008** | 3.13 | 4.60 | 4.53 | 6.81 | 5.11 |
| | (1.82) | (2.51) | (2.46) | (3.19) | (2.94) |
| **2010** | 3.14 | 4.75 | 4.80 | 7.47 | 5.49 |
| | (1.89) | (2.69) | (2.73) | (3.18) | (3.16) |
| **2012** | 3.25 | 4.54 | 5.34 | 8.23 | 5.84 |
| | (1.86) | (2.85) | (2.92) | (3.17) | (3.38) |
| **2014** | 3.74 | 5.05 | 5.74 | 8.95 | 6.36 |
| | (2.53) | (3.13) | (3.03) | (3.41) | (3.64) |
| **2016** | 4.32 | 5.43 | 6.81 | 9.96 | 7.19 |
| | (4.35) | (4.15) | (3.97) | (3.57) | (4.43) |
| **2018** | 4.20 | 5.57 | 7.45 | 10.36 | 7.51 |
| | (4.80) | (4.17) | (4.56) | (3.81) | (4.78) |
| **2020** | 3.47 | 6.67 | 8.40 | 11.78 | 8.46 |
| | (1.15) | (5.04) | (5.04) | (4.34) | (5.25) |
| **2022** | 3.61 | 6.27 | 8.02 | 10.93 | 8.04 |
| | (1.50) | (4.21) | (5.67) | (4.25) | (5.08) |
| **2024** | 3.28 | 6.30 | 8.51 | 11.25 | 8.23 |
| | (1.66) | (4.97) | (5.41) | (4.78) | (5.43) |

Calculated for countries with price data in real international dollar price data (177 to 181 countries over 9 periods)

**Supplemental Table S8: Mean real international dollar prices over time across World Bank 2024 income groups**

| | Low | Lower-middle | Upper-middle | High | All countries |
|---|---|---|---|---|---|
| **2008** | 2.69 | 4.05 | 4.39 | 6.64 | 4.53 |
| **2010** | 2.63 | 4.04 | 4.37 | 6.99 | 4.89 |
| **2012** | 2.64 | 3.97 | 5.17 | 7.78 | 5.25 |
| **2014** | 2.88 | 3.91 | 5.18 | 8.50 | 5.42 |
| **2016** | 3.12 | 4.42 | 6.35 | 9.23 | 6.41 |
| **2018** | 2.76 | 4.43 | 6.63 | 9.46 | 6.66 |
| **2020** | 3.77 | 5.48 | 7.39 | 10.89 | 7.60 |
| **2022** | 3.30 | 5.28 | 6.92 | 9.76 | 7.18 |
| **2024** | 3.35 | 5.05 | 7.48 | 10.04 | 7.44 |

Calculated for countries with price data in real international dollar price data (177 to 181 countries over 9 periods)

**Supplemental Table S9: Median annual consumer price inflation across regional and income groups, 2008-2024**

| | Income group in 2024 | | | | |
|---|---|---|---|---|---|
| **Region** | **Low** | **Lower-middle** | **Upper-middle** | **High** | **All** |
| AFRO | 5.9 | 5.6 | 4.3 | 2.6 | 5.3 |
| AMRO | | 5.7 | 3.6 | 2.4 | 3.6 |
| EMRO | 11.0 | 4.5 | 6.0 | 2.0 | 3.7 |
| EURO | | 8.5 | 5.2 | 2.1 | 2.6 |
| SEARO | | 6.3 | 2.6 | | 5.5 |
| WPRO | | 3.4 | 2.8 | 1.7 | 2.6 |
| **All** | 6.3 | 5.2 | 3.8 | 2.1 | 3.6 |

Note: Median CPI inflation computed over 396, 884, 881 and 1031 observations for low, lower-middle, upper-middle, and high income country (2024 World Bank classification) for the years 2008 to 2024. Venezuela dropped for computation of mean.

**Supplemental Table S10: Summary statistics of real local currency price changes (in %) over previous periods**

| | Lookback period (years) | | | | | | | |
|---|---|---|---|---|---|---|---|---|
| | **2** | **4** | **6** | **8** | **10** | **12** | **14** | **16** |
| **Mean** | 7% | 15% | 25% | 34% | 46% | 58% | 66% | 77% |
| **Median** | 3% | 10% | 17% | 23% | 31% | 37% | 40% | 49% |
| **75th percentile** | 14% | 26% | 39% | 54% | 67% | 81% | 90% | 101% |
| **Max** | 229% | 296% | 315% | 509% | 580% | 604% | 871% | 842% |
| **Min** | -72% | -75% | -69% | -80% | -72% | -70% | -69% | -67% |
| **N** | 1,372 | 1,199 | 1,025 | 855 | 678 | 507 | 339 | 170 |

Real price changes calculated as $(P_t - P_{t-x}/ P_t)$ ′ 100, where x = 2,4,…,16. Countries where real prices were over 200% higher than 10 years prior in one or more years included Peru (2018), the Bahamas, Iran, Qatar (in 2020), Montenegro, Saudi Arabia, Turkmenistan (2018, 2020), Ukraine(2018, 2024), Gambia (2018-22) and Philippines (2018-24)

Countries where real prices were over 200% higher than 16 years prior were the Bahamas, Bahrain, Belarus, Gambia, Maldives, Montenegro, Nicaragua, Oman, the Philippines, Qatar, Russian Federation, Saudi Arabi, Turkmenistan, Ukraine and Uzbekistan, all in the year 2024.

**Supplemental Table S11: Summary statistics of changes in real price in local currency over prior periods : Countries with price observations in all periods**

| | Periods prior to current period | | | | | | | |
|---|---|---|---|---|---|---|---|---|
| | **2** | **4** | **6** | **8** | **10** | **12** | **14** | **16** |
| **Mean** | 0.08 | 0.16 | 0.26 | 0.36 | 0.48 | 0.60 | 0.68 | 0.80 |
| **Median** | 0.03 | 0.10 | 0.18 | 0.24 | 0.32 | 0.38 | 0.40 | 0.50 |
| **75th percentile** | 0.14 | 0.26 | 0.40 | 0.55 | 0.68 | 0.81 | 0.90 | 1.01 |
| **Max** | 2.29 | 2.96 | 3.15 | 5.09 | 5.80 | 6.04 | 8.71 | 8.42 |
| **Min** | -0.72 | -0.71 | -0.68 | -0.69 | -0.68 | -0.64 | -0.69 | -0.67 |
| **N** | 1,250 | 1,092 | 934 | 777 | 620 | 464 | 309 | 154 |

**Supplemental Table S12: Summary statistics of changes in real price in local currency over prior periods : Countries with identical brands in all periods**

| | Periods prior to current period | | | | | | | |
|---|---|---|---|---|---|---|---|---|
| | **2** | **4** | **6** | **8** | **10** | **12** | **14** | **16** |
| **Mean** | 0.07 | 0.14 | 0.25 | 0.36 | 0.48 | 0.57 | 0.61 | 0.70 |
| **Median** | 0.03 | 0.09 | 0.17 | 0.24 | 0.32 | 0.38 | 0.37 | 0.41 |
| **75th percentile** | 0.13 | 0.27 | 0.44 | 0.59 | 0.68 | 0.84 | 0.82 | 0.92 |
| **Max** | 1.28 | 1.62 | 1.75 | 2.31 | 2.79 | 2.78 | 3.13 | 3.49 |
| **Min** | -0.41 | -0.40 | -0.48 | -0.54 | -0.60 | -0.49 | -0.53 | -0.61 |
| **N** | 481 | 420 | 359 | 298 | 237 | 177 | 118 | 59 |

**Supplemental Table S13: Share of countries with real local currency prices 50% over prior years by WHO region**

| vs years prior | 2 | 4 | 6 | 8 | 10 | 12 | 14 | 16 |
|---|---|---|---|---|---|---|---|---|
| AFRO | 7% | 11% | 24% | 28% | 28% | 37% | 39% | 39% |
| AMRO | 6% | 23% | 26% | 26% | 46% | 51% | 63% | 77% |
| EMRO | 25% | 20% | 35% | 25% | 60% | 55% | 70% | 70% |
| EURO | 4% | 4% | 13% | 17% | 34% | 51% | 68% | 77% |
| SEARO | 0% | 0% | 10% | 30% | 60% | 60% | 70% | 70% |
| WPRO | 4% | 13% | 13% | 17% | 39% | 39% | 57% | 57% |
| Total | **7%** | **12%** | **20%** | **23%** | **40%** | **47%** | **59%** | **64%** |

**Supplemental Table S14: Share of countries with real local prices 50% over prior years by 2024 World Bank income group**

| vs years prior | 2 | 4 | 6 | 8 | 10 | 12 | 14 | 16 |
|---|---|---|---|---|---|---|---|---|
| Low | 0.14 | 0.18 | 0.27 | 0.32 | 0.32 | 0.41 | 0.45 | 0.41 |
| Lower middle | 0.08 | 0.14 | 0.27 | 0.27 | 0.43 | 0.47 | 0.53 | 0.55 |
| Upper middle | 0.08 | 0.13 | 0.23 | 0.29 | 0.52 | 0.58 | 0.69 | 0.73 |
| High | 0.03 | 0.06 | 0.10 | 0.11 | 0.29 | 0.40 | 0.60 | 0.73 |
| **Total** | **0.07** | **0.12** | **0.20** | **0.23** | **0.40** | **0.47** | **0.59** | **0.64** |

**Supplemental Table S15: Annual rates of growth of real price in local currency units (2008-24) and implied time required to raise prices 50% and 20%**

| Country | Income group (2024) | Annual trend rate of growth of real price | | Implied time in full years to real price>1.5x | Implied time in full years to real price>1.2x |
|---|---|---|---|---|---|
| **AFRO** | | | | | |
| Algeria | Upper middle | 6.7% | ** | 7 | 3 |
| Angola | Lower middle | 4.5% | ** | 10 | 5 |
| Benin | Lower middle | -1.3% | ** | Flat/decline | Flat/decline |
| Botswana | Upper middle | 2.6% | ** | >10 | >5 |
| Burkina Faso | Low | 2.4% | ** | >10 | >5 |
| Burundi | Low | 1.0% | | >10 | >5 |
| Cabo Verde | Lower middle | 3.0% | ** | >10 | >5 |
| Cameroon | Lower middle | 0.7% | | >10 | >5 |
| Central African Republic | Low | -0.7% | | Flat/decline | Flat/decline |
| Chad | Low | -1.8% | ** | Flat/decline | Flat/decline |
| Comoros | Lower middle | -3.1% | * | Flat/decline | Flat/decline |
| Congo | Lower middle | 1.5% | ** | >10 | >5 |
| Côte d'Ivoire | Lower middle | -0.4% | | Flat/decline | Flat/decline |
| Democratic Republic of the Congo | Low | -6.5% | ** | Flat/decline | Flat/decline |
| Equatorial Guinea | Upper middle | -2.7% | ** | Flat/decline | Flat/decline |
| Eritrea | Low | | | | |
| Eswatini | Lower middle | -1.1% | ** | Flat/decline | Flat/decline |
| Ethiopia | Low | 3.2% | | >10 | >5 |
| Gabon | Upper middle | 0.1% | | >10 | >5 |
| Gambia | Low | 11.3% | ** | 4 | 2 |
| Ghana | Lower middle | 2.4% | | >10 | >5 |
| Guinea | Lower middle | -2.9% | ** | Flat/decline | Flat/decline |
| Guinea-Bissau | Low | 1.5% | | >10 | >5 |
| Kenya | Lower middle | 4.1% | * | >10 | 5 |
| Lesotho | Lower middle | 0.4% | | >10 | >5 |
| Liberia | Low | 2.0% | | >10 | >5 |
| Madagascar | Low | 0.9% | | >10 | >5 |
| Malawi | Low | -3.6% | | Flat/decline | Flat/decline |
| Mali | Low | -0.6% | | Flat/decline | Flat/decline |
| Mauritania | Lower middle | -3.1% | ** | Flat/decline | Flat/decline |
| Mauritius | Upper middle | 3.5% | ** | >10 | >5 |

| | | | | | |
|---|---|---|---|---|---|
| Mozambique | Low | 4.7% | ** | 9 | 4 |
| Namibia | Upper middle | 0.8% | | >10 | >5 |
| Niger | Low | 0.1% | | >10 | >5 |
| Nigeria | Lower middle | -4.2% | ** | Flat/decline | Flat/decline |
| Rwanda | Low | 1.4% | | >10 | >5 |
| Sao Tome and Principe | Lower middle | -4.6% | ** | Flat/decline | Flat/decline |
| Senegal | Lower middle | 27.4% | * | 2 | 1 |
| Seychelles | High | 24.5% | | 2 | 1 |
| Sierra Leone | Low | -0.4% | | Flat/decline | Flat/decline |
| South Africa | Upper middle | 23.5% | | 2 | 1 |
| South Sudan | Low | -18.6% | ** | Flat/decline | Flat/decline |
| Togo | Low | 2.6% | ** | >10 | >5 |
| Uganda | Low | 3.9% | ** | >10 | 5 |
| United Republic of Tanzania | Lower middle | -0.2% | | Flat/decline | Flat/decline |
| Zambia | Lower middle | 0.7% | | >10 | >5 |
| Zimbabwe | Lower middle | | | | |
| **AMRO** | | | | | |
| Antigua and Barbuda | High income | 2.2% | * | >10 | >5 |
| Argentina | Upper middle income | -0.2% | | Flat/decline | Flat/decline |
| Bahamas | High income | 8.5% | ** | 5 | 3 |
| Barbados | High income | 0.7% | | >10 | >5 |
| Belize | Upper middle income | 0.5% | | >10 | >5 |
| Bolivia (Plurinational State of) | Lower middle income | 2.2% | ** | >10 | >5 |
| Brazil | Upper middle income | 0.1% | | >10 | >5 |
| Canada | High income | 1.8% | ** | >10 | >5 |
| Chile | High income | 4.7% | ** | 9 | 5 |
| Colombia | Upper middle income | 6.1% | ** | 7 | 4 |
| Costa Rica | Upper middle income | 4.2% | ** | 10 | 5 |
| Cuba | Upper middle income | | | | |
| Dominica | Upper middle income | 4.0% | ** | >10 | 5 |
| Dominican Republic | Upper middle income | 4.3% | ** | 10 | 5 |
| Ecuador | Upper middle income | 7.1% | ** | 6 | 3 |
| El Salvador | Upper middle income | 4.4% | ** | 10 | 5 |
| Grenada | Upper middle income | 2.0% | * | >10 | >5 |

| | | | | | |
|---|---|---|---|---|---|
| Guatemala | Upper middle income | 0.3% | | >10 | >5 |
| Guyana | High income | 2.4% | ** | >10 | >5 |
| Haiti | Lower middle income | -2.1% | ** | Not enough years | Not enough years |
| Honduras | Lower middle income | 4.0% | ** | >10 | 5 |
| Jamaica | Upper middle income | 2.0% | ** | >10 | >5 |
| Mexico | Upper middle income | 2.3% | ** | >10 | >5 |
| Nicaragua | Lower middle income | 7.3% | ** | 6 | 3 |
| Panama | High income | 3.3% | ** | >10 | >5 |
| Paraguay | Upper middle income | 3.0% | ** | >10 | >5 |
| Peru | Upper middle income | 6.3% | ** | 7 | 4 |
| Saint Kitts and Nevis | High income | 3.1% | ** | >10 | >5 |
| Saint Lucia | Upper middle income | 3.7% | ** | >10 | >5 |
| Saint Vincent and the Grenadines | Upper middle income | 2.4% | * | >10 | >5 |
| Suriname | Upper middle income | 3.4% | ** | >10 | >5 |
| Trinidad and Tobago | High income | 0.4% | | >10 | >5 |
| United States of America | High income | 1.3% | ** | >10 | >5 |
| Uruguay | High income | 1.7% | ** | >10 | >5 |
| Venezuela (Bolivarian Republic of) | Lower middle income | | | | |
| **EMRO** | | | | | |
| Afghanistan | Low income | | | | |
| Bahrain | High income | 8.1% | ** | 6 | 3 |
| Djibouti | Lower middle income | -0.4% | | Flat/decline | Flat/decline |
| Egypt | Lower middle income | 3.3% | ** | >10 | >5 |
| Iran (Islamic Republic of) | Upper middle income | 8.1% | ** | 6 | 3 |
| Iraq | Upper middle income | 3.6% | | >10 | >5 |
| Jordan | Lower middle income | 5.1% | ** | 9 | 4 |
| Kuwait | High income | -0.9% | | Flat/decline | Flat/decline |
| Lebanon | Lower middle income | -7.2% | ** | Flat/decline | Flat/decline |
| Libya | Upper middle income | -0.1% | | Flat/decline | Flat/decline |

| | | | | | |
|---|---|---|---|---|---|
| Morocco | Lower middle income | 1.3% | ** | >10 | >5 |
| Oman | High income | | | | |
| Pakistan | Lower middle income | 8.2% | ** | 6 | 3 |
| Qatar | High income | 4.5% | ** | 10 | 5 |
| Saudi Arabia | High income | 8.3% | ** | 6 | 3 |
| Somalia | Low income | 9.3% | ** | 5 | 3 |
| Sudan | Low income | -4.3% | ** | Flat/decline | Flat/decline |
| Syrian Arab Republic | Low income | -8.7% | ** | Flat/decline | Flat/decline |
| Tunisia | Lower middle income | | | | |
| United Arab Emirates | High income | 0.7% | * | >10 | >5 |
| occupied Palestinian territory | Lower middle income | 7.9% | ** | 6 | 3 |
| Yemen | Low income | -2.6% | ** | Flat/decline | Flat/decline |
| **EURO** | | | | | |
| Albania | Upper middle income | 2.6% | ** | >10 | >5 |
| Andorra | High income | 1.8% | * | >10 | >5 |
| Armenia | Upper middle income | 0.4% | | >10 | >5 |
| Austria | High income | 1.1% | ** | >10 | >5 |
| Azerbaijan | Upper middle income | 2.5% | | >10 | >5 |
| Belarus | Upper middle income | 8.1% | ** | 6 | 3 |
| Belgium | High income | 3.0% | ** | >10 | >5 |
| Bosnia and Herzegovina | Upper middle income | 6.5% | ** | 7 | 3 |
| Bulgaria | High income | 0.8% | | >10 | >5 |
| Croatia | High income | 0.8% | | >10 | >5 |
| Cyprus | High income | 2.2% | ** | >10 | >5 |
| Czechia | High income | 3.4% | ** | >10 | >5 |
| Denmark | High income | 2.8% | ** | >10 | >5 |
| Estonia | High income | 2.0% | * | >10 | >5 |
| Finland | High income | 3.9% | ** | >10 | 5 |
| France | High income | 3.7% | ** | >10 | 5 |
| Georgia | Upper middle income | 5.8% | ** | 8 | 4 |
| Germany | High income | 1.8% | ** | >10 | >5 |
| Greece | High income | 1.7% | ** | >10 | >5 |
| Hungary | High income | 5.3% | ** | 8 | 4 |
| Iceland | High income | 1.5% | ** | >10 | >5 |
| Ireland | High income | 3.9% | ** | >10 | 5 |
| Israel | High income | 3.3% | ** | >10 | >5 |

| | | | | | |
|---|---|---|---|---|---|
| Italy | High income | 1.7% | ** | >10 | >5 |
| Kazakhstan | Upper middle income | 6.9% | ** | 7 | 3 |
| Kyrgyzstan | Lower middle income | 6.8% | ** | 7 | 3 |
| Latvia | High income | 2.5% | ** | >10 | >5 |
| Lithuania | High income | 4.0% | ** | >10 | 5 |
| Luxembourg | High income | 1.7% | ** | >10 | >5 |
| Malta | High income | 1.1% | * | >10 | >5 |
| Monaco | High income | | | | |
| Montenegro | Upper middle income | 9.2% | ** | 5 | 3 |
| Netherlands (Kingdom of the) | High income | 2.9% | ** | >10 | >5 |
| North Macedonia | Upper middle income | 3.8% | ** | >10 | 5 |
| Norway | High income | 2.7% | ** | >10 | >5 |
| Poland | High income | 3.7% | ** | >10 | >5 |
| Portugal | High income | 0.7% | | >10 | >5 |
| Republic of Moldova | Upper middle income | 6.2% | ** | 7 | 4 |
| Romania | High income | 4.2% | ** | 10 | 5 |
| Russian Federation | High income | 7.5% | ** | 6 | 3 |
| San Marino | High income | 1.1% | ** | >10 | >5 |
| Serbia | Upper middle income | 6.5% | ** | 7 | 3 |
| Slovakia | High income | 2.6% | ** | >10 | >5 |
| Slovenia | High income | 2.9% | ** | >10 | >5 |
| Spain | High income | 2.4% | * | >10 | >5 |
| Sweden | High income | 1.1% | ** | >10 | >5 |
| Switzerland | High income | 1.6% | ** | >10 | >5 |
| Tajikistan | Lower middle income | 4.8% | ** | 9 | 4 |
| Türkiye | Upper middle income | 2.7% | * | >10 | >5 |
| Turkmenistan | Upper middle income | 13.6% | ** | 4 | 2 |
| Ukraine | Upper middle income | 11.3% | ** | 4 | 2 |
| United Kingdom | High income | 2.2% | ** | >10 | >5 |
| Uzbekistan | Lower middle income | 8.6% | ** | 5 | 3 |
| **SEARO** | | | | | |
| Bangladesh | Lower middle income | 7.3% | ** | 6 | 3 |
| Bhutan | Lower middle income | -15.7% | | Not enough years | Not enough years |

| | | | | | |
|---|---|---|---|---|---|
| Democratic People's Republic of Korea | Low income | | | | |
| India | Lower middle income | 1.9% | | >10 | >5 |
| Indonesia | Upper middle income | 3.6% | ** | >10 | >5 |
| Maldives | Upper middle income | 9.3% | ** | 5 | 3 |
| Myanmar | Lower middle income | -3.2% | * | Flat/decline | Flat/decline |
| Nepal | Lower middle income | 4.4% | ** | 10 | 5 |
| Sri Lanka | Lower middle income | 6.2% | ** | 7 | 4 |
| Thailand | Upper middle income | 0.5% | | >10 | >5 |
| Timor-Leste | Lower middle income | 0.6% | | >10 | >5 |
| **WPRO** | | | | | |
| Australia | High income | 6.2% | ** | 7 | 4 |
| Brunei Darussalam | High income | 29.5% | | Not enough years | Not enough years |
| Cambodia | Lower middle income | 0.5% | | >10 | >5 |
| China | Upper middle income | 4.1% | ** | >10 | 5 |
| Cook Islands | High income | | | | |
| Fiji | Upper middle income | 7.1% | ** | 6 | 3 |
| Japan | High income | 3.7% | ** | >10 | 5 |
| Kiribati | Lower middle income | 0.2% | | >10 | >5 |
| Lao People's Democratic Republic | Lower middle income | -2.4% | * | Flat/decline | Flat/decline |
| Malaysia | Upper middle income | 2.9% | ** | >10 | >5 |
| Marshall Islands | Upper middle income | 0.4% | | >10 | >5 |
| Micronesia (Federated States of) | Lower middle income | 6.0% | ** | 7 | 4 |
| Mongolia | Upper middle income | 2.5% | | >10 | >5 |
| Nauru | High income | 6.8% | ** | 7 | 3 |
| New Zealand | High income | 6.7% | ** | 7 | 3 |
| Niue | High income | | | | |
| Palau | High income | 2.6% | * | >10 | >5 |
| Papua New Guinea | Lower middle income | -0.3% | | Flat/decline | Flat/decline |
| Philippines | Lower middle income | 14.8% | ** | 3 | 2 |

| | | | | | |
|---|---|---|---|---|---|
| Republic of Korea | High income | 3.0% | ** | >10 | >5 |
| Samoa | Lower middle income | 2.5% | ** | >10 | >5 |
| Singapore | High income | 0.2% | | >10 | >5 |
| Solomon Islands | Lower middle income | -0.8% | | Flat/decline | Flat/decline |
| Tonga | Upper middle income | 5.9% | ** | 8 | 4 |
| Tuvalu | Upper middle income | 2.0% | | >10 | >5 |
| Vanuatu | Lower middle income | -0.6% | * | Flat/decline | Flat/decline |
| Viet Nam | Lower middle income | -0.5% | | Flat/decline | Flat/decline |

Note: Growth rates are the coefficients of a simple linear regression of the natural log of each country’s real price on year and a constant. Number of periods of data varies from 5 to 9 years. Growth rates were not calculated for observations with fewer than 4 years of data

**Supplemental Table S16: Percent increase in price over previous period by period-specific deciles of percent increase in excise tax, local currency units**

| Deciles of % excise change | Mean % increase in LCU price over 2 years prior | | | | | | | | Summary statistics, pooled | | | |
|---|---|---|---|---|---|---|---|---|---|---|---|---|
| | **2010** | **2012** | **2014** | **2016** | **2018** | **2020** | **2022** | **2024** | **Mean** | **Median** | **Min** | **Max** |
| **1** | 9% | 5% | 4% | 0% | -5% | 2% | 3% | 2% | 3% | 0% | -100% | 95% |
| **2** | | 10% | 4% | | 9% | 0% | 10% | | 7% | 0% | -89% | 95% |
| **3** | 41% | 6% | 10% | 5% | 0% | -4% | 1% | 5% | 9% | 2% | -68% | 98% |
| **4** | 23% | 8% | -2% | 6% | 8% | 5% | 5% | 6% | 7% | 6% | -140% | 98% |
| **5** | 28% | 12% | 10% | 8% | 6% | 7% | 3% | 11% | 10% | 8% | -20% | 98% |
| **6** | 28% | 14% | 15% | 10% | 10% | 10% | 15% | 17% | 15% | 11% | -9% | 98% |
| **7** | 23% | 2% | 9% | 19% | 15% | 13% | 14% | 14% | 14% | 15% | -96% | 99% |
| **8** | 23% | 20% | 21% | 22% | 12% | 15% | 19% | 18% | 19% | 20% | -88% | 99% |
| **9** | 26% | 31% | 20% | 23% | 24% | 30% | 23% | 34% | 26% | 28% | -54% | 96% |
| **10** | 28% | 33% | 33% | 30% | 28% | 48% | 30% | 51% | 35% | 37% | -133% | 100% |
| All deciles | 23% | 14% | 12% | 12% | 11% | 13% | 13% | 16% | 14% | 11% | -140% | 100% |
| N | 157 | 132 | 162 | 164 | 163 | 163 | 162 | 172 | 1,275 | | | |

Note: Excise tax increases are computed as the increase in excise tax yield (i.e. amount of tax per 20 cigarettes) between successive GTCR reports. Outliers defined as instances of local currency price change of less than -200%, as a result, 40 observations were dropped.

**Supplemental Table S17: % increase in excise and tax regime type by price increase**

| | % change in excise between 2 years | | | | | Tax regime | | | | Count |
|---|---|---|---|---|---|---|---|---|---|---|
| **% price change** | **Mean** | **SD** | **Median** | **Min** | **Max** | **Specific** | **Ad val** | **Mixed** | **No excise** | |
| less than 0 | -8% | 41% | -3% | -291% | 100% | 33% | **39%** | 18% | 9% | 293 |
| 0 to 10pc | 8% | 14% | 5% | -43% | 100% | 35% | 18% | **42%** | 5% | 449 |
| 10 to 20pc | 14% | 17% | 13% | -126% | 100% | **40%** | 13% | **41%** | 7% | 309 |
| 20 to 30pc | 28% | 25% | 25% | -51% | 100% | 36% | 24% | 31% | 10% | 137 |
| 30 to 40pc | 30% | 38% | 33% | -227% | 100% | **42%** | 21% | 26% | 11% | 68 |
| 40 to 50pc | 44% | 30% | 45% | -6% | 100% | **52%** | 15% | 18% | 15% | 28 |
| 50pc and over | 66% | 31% | 70% | -14% | 100% | 29% | **35%** | 19% | 17% | 12 |
| All | 10% | 30% | 8% | -291% | 100% | **35%** | 25% | 31% | 9% | 1,296 |

Note: USD price changes greater than -200% only (4 observations dropped). Pooled over 8 periods (even numbered years 2010-24).

**Supplemental Table S18: Excise tax changes associated with 2-year PPP$ price increases of over 50%: Instances of two-year increase over 50% and explanations**

| Region | Year | Country | PPP$ price change, % | PPP$ excise change, % | Excise in LCU, 2 years prior | Excise in LCU, current | Explanation |
|---|---|---|---|---|---|---|---|
| AFRO | 2014 | Algeria | 78% | 1% | 21.42 | 21.42 | Most-sold brand price increase without tax increase |
| AFRO | 2014 | Burundi | 82% | 30% | 307.49 | 440.00 | Specific tax introduced |
| AFRO | 2014 | Ethiopia | 66% | -9% | 2.03 | 2.09 | Most-sold brand price increase without tax increase |
| AFRO | 2014 | Gambia | 86% | 153% | 3.30 | 9.00 | Specific tax ~ tripled, 3.3 to 9 GMD/20 sticks |
| AFRO | 2020 | Ethiopia | 111% | 518% | 2.09 | 16.31 | Specific tax introduced, 8 ETB/20 sticks |
| AFRO | 2020 | Kenya | 89% | 24% | 50.00 | 63.14 | Specific tax increased, 2500 to 3157 KES |
| AFRO | 2020 | Liberia | 79% | 111% | 28.95 | 79.76 | Specific tax introduced, 79.76 LRD/20 sticks |
| AFRO | 2024 | Ethiopia | 93% | 79% | 14.18 | 39.46 | Specific tax increased, 8 to 20 ETB/20 sticks |
| AFRO | 2024 | Gabon | 127% | 79% | 197.05 | 311.74 | Specific tax increased, 150 to 250 XAF |
| AFRO | 2024 | Ghana | 75% | 302% | 1.08 | 7.15 | Specific tax introduced, 0.28 GHS/stick |
| AFRO | 2024 | Zambia | 67% | -6% | 1.78 | 2.00 | Specific tax increased, 88.75 to 100 ZMW/ 1000 sticks |
| AMRO | 2010 | Panama | 55% | 235% | 0.42 | 1.50 | Specific tax introduced, 1.5 USD/20 sticks |
| AMRO | 2012 | Bahamas | 89% | 60% | 0.84 | 1.32 | Most-sold brand price increase without tax increase |
| AMRO | 2012 | Iran (Islamic Republic of) | 76% | . | 0.00 | 0.00 | Most-sold brand price increase without tax increase |
| AMRO | 2016 | Ecuador | 79% | 84% | 1.85 | 3.20 | Specific tax increased, 0.0925 to 0.16 USD/ stick |
| AMRO | 2016 | Peru | 62% | 146% | 1.40 | 3.60 | Specific tax increased, 0.07 to 0.18 PEN/stick |
| AMRO | 2018 | Saint Vincent and the Grenadines | 70% | 2% | 0.31 | 0.31 | Most-sold brand price increase without tax increase |
| AMRO | 2020 | Antigua and Barbuda | 91% | . | 0.00 | 0.00 | Most-sold brand price increase without tax increase |

| | | | | | | | |
|---|---|---|---|---|---|---|---|
| AMRO | 2020 | Nicaragua | 153% | 318% | 12.21 | 50.00 | Specific tax increased, 610.53 to 2500 NIO/1000 sticks |
| AMRO | 2024 | Saint Lucia | 65% | 111% | 3.52 | 7.12 | Specific tax increased, 176 to 356 NIO/1000 sticks |
| EMRO | 2010 | Turkmenistan | 64% | 190% | 2600.05 | 1.43 | Minimum specific tax increased |
| EMRO | 2014 | Libya | 109% | . | 0.00 | 0.00 | Most-sold brand price increase without tax increase |
| EMRO | 2018 | Iran (Islamic Republic of) | 74% | 49% | 2000.00 | 4300.00 | Most-sold brand price increase without tax increase |
| EMRO | 2018 | Saudi Arabia | 100% | . | 0.00 | 13.41 | Ad valorem tax introduced, 100% of pre-tax price |
| EMRO | 2018 | United Arab Emirates | 80% | . | 0.00 | 9.76 | Ad valorem tax introduced, 100% of pre-tax price |
| EMRO | 2020 | Iran (Islamic Republic of) | 105% | 99% | 4300.00 | 16655.00 | Most-sold brand price increase without tax increase |
| EMRO | 2020 | Oman | 102% | . | 0.00 | 1.10 | Ad valorem tax introduced, 100% of pre-tax price |
| EMRO | 2020 | Qatar | 126% | . | 0.00 | 11.00 | Ad valorem tax introduced, 100% of pre-tax price |
| EMRO | 2024 | Iraq | 232% | . | 0.00 | 0.00 | Most-sold brand price increase without tax increase |
| EURO | 2010 | Ukraine | 75% | 191% | 0.80 | 2.95 | Specific tax increased, 30 to 90 UKH/1000 sticks |
| EURO | 2012 | Maldives | 68% | . | 0.00 | 0.00 | Import duty increased, 30 to 90 MVR/stick |
| EURO | 2012 | Turkmenistan | 133% | -12% | 1.43 | 1.43 | Minimum specific tax increased |
| EURO | 2014 | Belarus | 68% | 216% | 576.00 | 2500.00 | Specific tax increased, 28800 to 12500 BYR/1000 sticks |
| EURO | 2016 | Azerbaijan | 95% | 176% | 0.03 | 0.08 | Specific tax increased, 1.4112 to 4 AZN/1000 sticks |
| EURO | 2018 | Ukraine | 49% | 37% | 8.51 | 15.46 | Minimum specific tax increased |
| EURO | 2024 | Belarus | 108% | 214% | 0.66 | 2.14 | Specific tax increased, 32.94 to 106.88 BYN/1000 sticks |
| SEARO | 2012 | Brunei | 182% | 256% | 1.20 | 5.00 | Specific tax increased, 60 to 250 BND/1000 sticks |

| | | | | | | | |
|---|---|---|---|---|---|---|---|
| SEARO | 2016 | Sri Lanka | 56% | 34% | 332.20 | 475.00 | Specific tax increased, 16610 to 23750 LKR/1000 sticks |
| SEARO | 2024 | Sri Lanka | 64% | 55% | 862.00 | 1529.80 | Specific tax increased, 43100 to 76490 LKR/1000 sticks |
| WPRO | 2014 | Mongolia | 87% | 161% | 322.14 | 898.04 | Specific tax increased, 2 to 2.5 USD/ 100 cigarettes |
| WPRO | 2014 | Philippines | 75% | 515% | 2.72 | 17.00 | Specific tax increased, 2.72 to 17 PHP/20 sticks |
| WPRO | 2016 | Republic of Korea | 74% | 113% | 1322.50 | 2914.00 | Specific tax increased, 1322.5 to 2914 KRW/20 sticks |
| WPRO | 2018 | Tuvalu | 62% | -69% | 1.50 | 0.49 | Most-sold brand price increase without tax increase |
| WPRO | 2020 | Philippines | 78% | 32% | 35.00 | 45.00 | Specific tax increased, 35 to 45 PHP/20 sticks |
| WPRO | 2022 | Tuvalu | 103% | 1278% | 0.15 | 2.24 | Ad valorem tax increased |
| WPRO | 2024 | Kiribati | 91% | -4% | 2.20 | 2.20 | Most-sold brand price increase without tax increase |
| WPRO | 2024 | Marshall Islands | 89% | . | 0.00 | 0.00 | Most-sold brand price increase without tax increase |

**Supplemental Table S19: Drivers of price increase in countries with 50% or larger price increase over 2 year windows**

| Driver of price increase | Count |
|---|---|
| New ad val tax | 4 |
| New specific tax | 6 |
| Ad val rate increase | 1 |
| Specific rate or minimum specific tax increase | 23 |
| Import duty increased | 1 |
| Price increase without tax increase | 13 |
| All | 48 |

**Supplemental Table S20: Pass-through of excise tax to prices: Coefficients from two-way fixed-effects regression of ln(price of cigarettes in international $) on ln(Excise Tax in international $)**

| | **Full sample** | | | | **High income, 2024** | | **LMIC, 2024** | |
|---|---|---|---|---|---|---|---|---|
| | **(I)** | | **(II)** | | **(III)** | | **(IV)** | |
| ln (excise tax, PPP$) | 0.32 | ** | 0.33 | ** | 0.38 | * | 0.32 | ** |
| | (0.04) | 0 | (0.04) | 0 | (0.15) | 0.2 | (0.04) | 0 |
| | 0.045022 | | 0.045368 | | 0.050943 | | 0.044198 | |
| Constant | 1.19 | ** | 1.21 | ** | 1.22 | ** | 1.15 | ** |
| | (0.02) | 0 | (0.04) | 0 | (0.13) | 0.1 | (0.03) | 0 |
| | 0.22 | | 0.43 | | 0.13 | | 0.28 | |
| Mixed | | | -0.06 | | | | | |
| Specific | | | (0.06) | 0.1 | | | | |
| | | | 0.00 | | | | | |
| | | | (0.06) | 0.1 | | | | |
| Overall R-squared | 0.73 | | 0.73 | | 0.74 | | 0.64 | |
| # countries | 181 | | 181 | | 59 | | 122 | |
| N (observations) | 1485 | | 1485 | | 481 | | 1004 | |

Note: Panel with observations in even-numbered years, 2008-24. Robust standard errors (clustered at the country level)

*** = significant at 99%, **= significant at 95%, * = significant at 90%

Omitted tax regime category = ad valorem. Restricted to observations where the dependent variable, 2-year price changes, is greater than -500%

**Supplemental Table S21: Interaction of price measures and tax regime**

| | (I) | | | (II) | |
|---|---|---|---|---|---|
| Excise in PPP$ | 1.53 | *** | ln (excise tax, PPP$) | 0.26 | *** |
| | (0.18) | 0.180605 | | (0.04) | 0.150109 |
| | 0.045022 | | | 0.050943 | |
| Constant | 1.21 | *** | Constant | 1.15 | *** |
| | (0.42) | 0.427199 | | (0.03) | 0.124473 |
| | 0.22 | | | 0.13 | |
| Mixed | 0.40 | | Mixed | -0.01 | |
| | (0.38) | 0.38 | | (0.06) | 0.06 |
| Specific | 0.38 | | Specific | 0.05 | |
| | -0.48 | | | (0.05) | 0.05 |
| Excise in PPP$ x Mixed | -0.23 | | ln(Excise in PPP$) x Mixed | 0.13 | *** |
| | (0.16) | 0.159395 | | (0.04) | ** |
| Excise in PPP$ x Specific | 0.01 | | ln(Excise in PPP$) x Specific | 0.10 | |
| | (0.17) | | | (0.05) | |
| Overall R-squared | 0.79 | | | 0.79 | |
| # countries | 181 | | | 59 | |
| N (observations) | 1486 | | | 1489 | |

Note: Panel with observations in even-numbered years, 2008-24. Robust standard errors (clustered at the country level)

*** = significant at 99%, **= significant at 95%, * = significant at 90%

Omitted tax regime category = ad valorem. Restricted to observations where the dependent variable, 2-year price changes, is greater than -500%

**Supplemental Table S22:Terminal digits in nominal local currency prices**

| | Fraction of price observations ending in "..0.00" | | | | | | | | | |
|---|---|---|---|---|---|---|---|---|---|---|
| **Region** | **2008** | **2010** | **2012** | **2014** | **2016** | **2018** | **2020** | **2022** | **2024** | **All years** |
| AFRO | 0.69 | 0.70 | 0.67 | 0.77 | 0.76 | 0.74 | 0.86 | 0.88 | 0.84 | 0.77 |
| AMRO | 0.15 | 0.24 | 0.21 | 0.21 | 0.23 | 0.39 | 0.23 | 0.31 | 0.29 | 0.25 |
| EMRO | 0.23 | 0.27 | 0.32 | 0.45 | 0.50 | 0.48 | 0.43 | 0.33 | 0.45 | 0.38 |
| EURO | 0.10 | 0.11 | 0.12 | 0.15 | 0.13 | 0.10 | 0.15 | 0.15 | 0.13 | 0.13 |
| SEARO | 0.40 | 0.33 | 0.22 | 0.30 | 0.40 | 0.50 | 0.64 | 0.64 | 0.70 | 0.47 |
| WPRO | 0.27 | 0.33 | 0.31 | 0.28 | 0.27 | 0.27 | 0.33 | 0.32 | 0.30 | 0.30 |
| **All regions** | 0.31 | 0.33 | 0.32 | 0.37 | 0.37 | 0.39 | 0.42 | 0.42 | 0.42 | 0.37 |

| | Fraction of price observations ending in "..5.00" | | | | | | | | | |
|---|---|---|---|---|---|---|---|---|---|---|
| **Region** | **2008** | **2010** | **2012** | **2014** | **2016** | **2018** | **2020** | **2022** | **2024** | **All years** |
| AFRO | 0.09 | 0.05 | 0.16 | 0.09 | 0.07 | 0.09 | 0.02 | 0.02 | 0.02 | 0.07 |
| AMRO | 0.12 | 0.03 | 0.12 | 0.06 | 0.06 | 0.06 | 0.13 | 0.06 | 0.06 | 0.08 |
| EMRO | 0.00 | 0.05 | 0.05 | 0.00 | 0.00 | 0.00 | 0.00 | 0.06 | 0.05 | 0.02 |
| EURO | 0.04 | 0.02 | 0.02 | 0.10 | 0.06 | 0.12 | 0.08 | 0.04 | 0.08 | 0.06 |
| SEARO | 0.30 | 0.22 | 0.11 | 0.10 | 0.00 | 0.00 | 0.18 | 0.09 | 0.00 | 0.11 |
| WPRO | 0.00 | 0.00 | 0.04 | 0.00 | 0.00 | 0.04 | 0.04 | 0.08 | 0.09 | 0.03 |
| **All regions** | 0.07 | 0.04 | 0.08 | 0.06 | 0.04 | 0.07 | 0.07 | 0.05 | 0.05 | 0.06 |

**Supplemental Figure F1: Change in real local currency prices over previous period by WHO region (countries with 4 or more price observations)**

**AFRO**

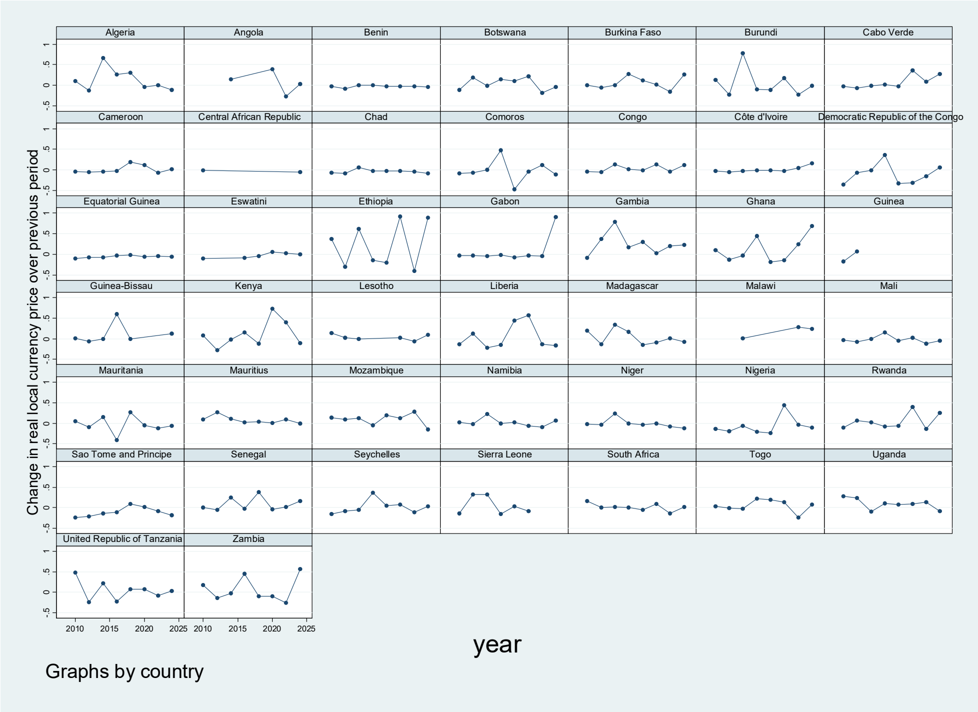

**AMRO**

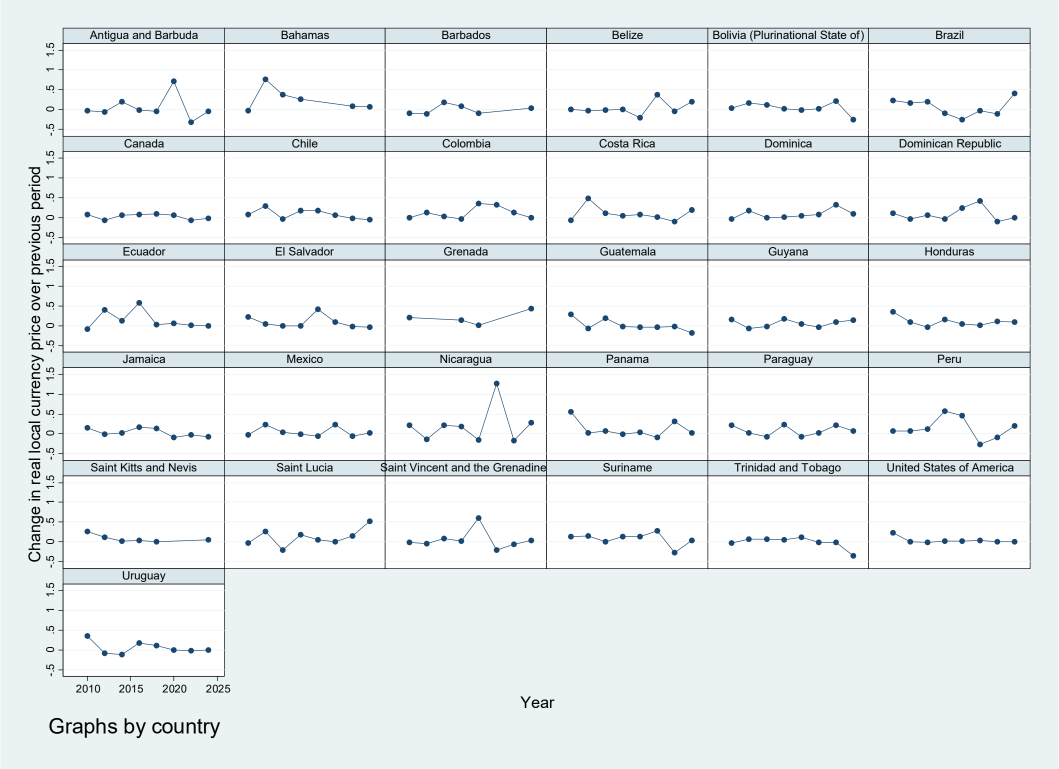

**EMRO**

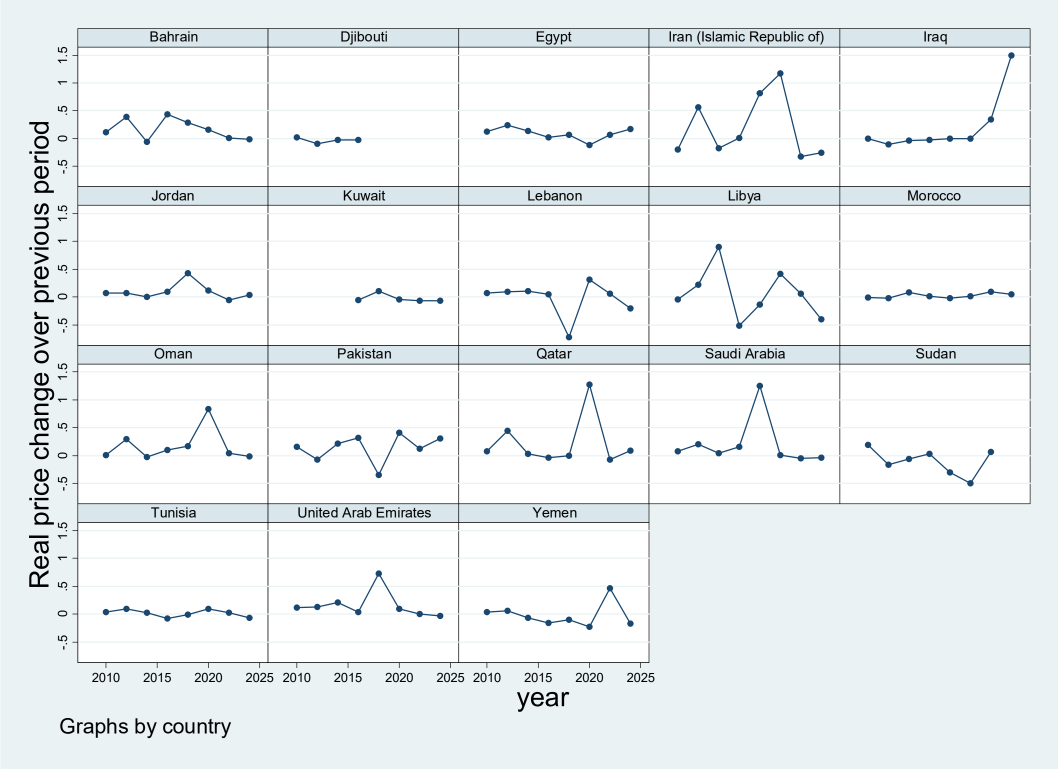

Bahrain
Djibouti
Egypt
Iran (Islamic Republic of)
Iraq
Jordan
Kuwait
Lebanon
Libya
Morocco
Oman
Pakistan
Qatar
Saudi Arabia
Sudan
Tunisia
United Arab Emirates
Yemen
Real price change over previous period
-.5
0
.5
1
1.5
2010
2015
2020
2025
year
Graphs by country

**EURO**

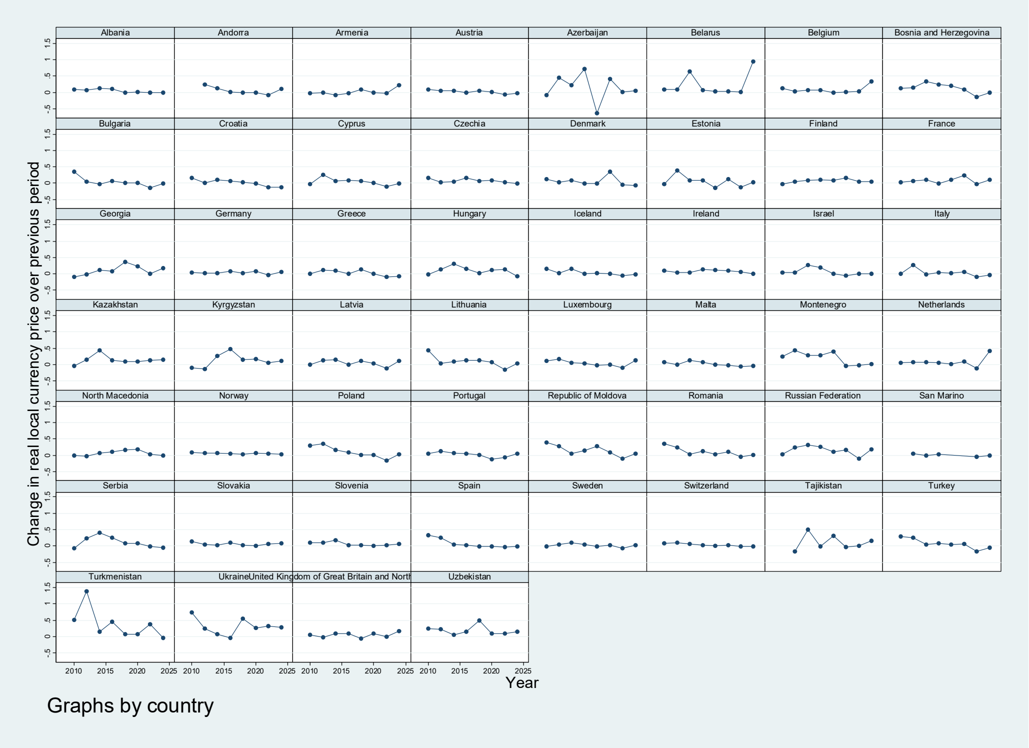

Albania
Andorra
Armenia
Austria
Azerbaijan
Belarus
Belgium
Bosnia and Herzegovina
Bulgaria
Croatia
Cyprus
Czechia
Denmark
Estonia
Finland
France
Georgia
Germany
Greece
Hungary
Iceland
Ireland
Israel
Italy
Kazakhstan
Kyrgyzstan
Latvia
Lithuania
Luxembourg
Malta
Montenegro
Netherlands
North Macedonia
Norway
Poland
Portugal
Republic of Moldova
Romania
Russian Federation
San Marino
Serbia
Slovakia
Slovenia
Spain
Sweden
Switzerland
Tajikistan
Turkey
Turkmenistan
UkraineUnited Kingdom of Great Britain and North
Uzbekistan
Change in real local currency price over previous period
2010
2015
2020
2025
Year
Graphs by country

**SEARO**

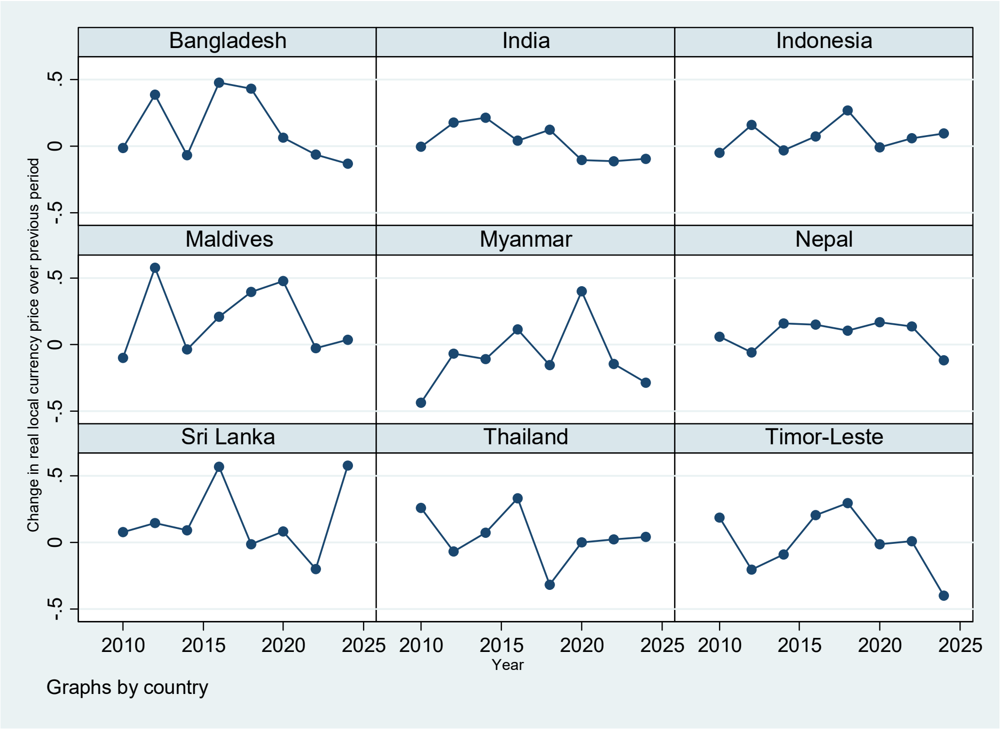
Bangladesh
India
Indonesia
Maldives
Myanmar
Nepal
Sri Lanka
Thailand
Timor-Leste
Change in real local currency price over previous period
-.5
0
.5
2010
2015
2020
2025
Year
Graphs by country

**WPRO**

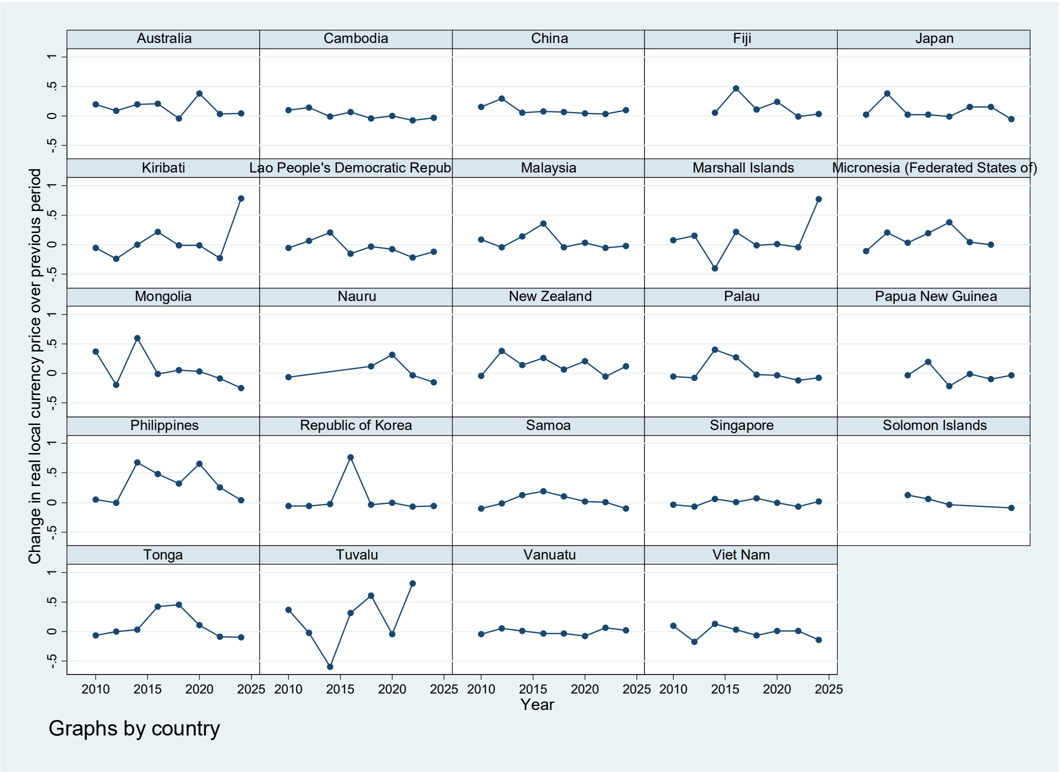
Australia
Cambodia
China
Fiji
Japan
Kiribati
Lao People's Democratic Repub
Malaysia
Marshall Islands
Micronesia (Federated States of)
Mongolia
Nauru
New Zealand
Palau
Papua New Guinea
Philippines
Republic of Korea
Samoa
Singapore
Solomon Islands
Tonga
Tuvalu
Vanuatu
Viet Nam
Change in real local currency price over previous period
-.5
0
.5
1
2010
2015
2020
2025
Year
Graphs by country

**Supplemental Figure F2: Distribution of real price changes over 2- and 10-year lookback period**

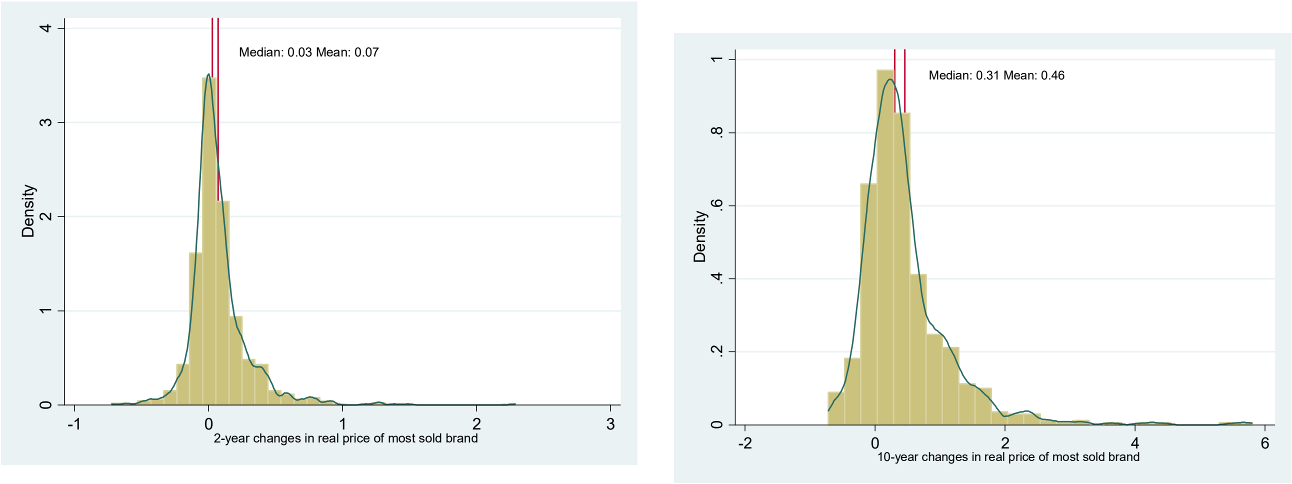

**Supplemental Figure F3: Fraction of countries with real prices 20% and 50% higher by price quintile**

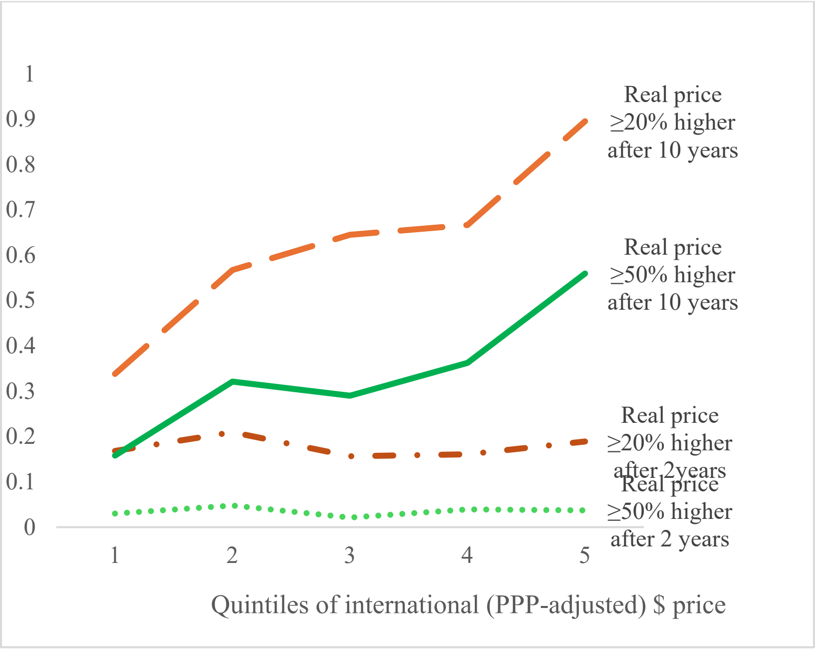

Note: Quintiles of international dollar price computed in each year. Years 2018-24 included for 10-year lookback, and years 2010-24 included for 2-year lookback. Country counts vary by year, with 33 to 36 countries per quintile in a given year.
